# Metabolites and lipoproteins may predict the severity of early Acute Pancreatitis in a South African cohort

**DOI:** 10.1101/2023.11.03.23298015

**Authors:** Jeanet Mazibuko, Nnenna Elebo, Aurelia A. Williams, Jones Omoshoro-Jones, John W. Devar, Martin Smith, Stefano Cacciatore, Pascaline N. Fru

## Abstract

**Background:** Acute pancreatitis (AP) is a common clinical disease with varying severity. The Revised Atlanta Classification (RAC) categorises AP into mild, moderately severe, and severe (MAP, MSAP and SAP) respectively. Despite the availability of different scoring systems to triage patients, these are not always suitable for predicting the course and outcome of certain patients during admission. In this study, untargeted metabolomics was used to identify metabolic parameters that can potentially be used as prognostic markers for stratifying the risk profiles of patients for improved management and treatment.

**Methods:** Serum isolated from blood samples collected from 30 AP patients (8 MAP, 14 MSAP, and 8 SAP) and 9 healthy control (HC) individuals was analysed using ^1^H nuclear magnetic resonance (NMR) spectroscopy. Wilcoxon and Kruskal–Walli’s rank-sum tests were used to compare differences in numerical covariates. A liposcale test was used for lipoprotein characterization and the Spearman rank test was conducted for correlation of the data. P-values < 0.05 were considered significant.

**Results:** Elevated levels of lactate, (rho = 0.67; p-value < 0.001, FDR = 0.001), 3-hydroxybutyrate (rho = 0.46; p-value < 0.003, FDR = 0.013), acetoacetate (rho = 0.63; p-value < 0.001, FDR < 0.001) and lipid alpha-CH2 (rho = 0.45; p-value = 0.004, FDR = 0.013) were associated with AP severity as was decreased levels of ascorbate (rho = 0.46; p-value < 0.003, FDR = 0.013), methanol (rho = 0.46; p-value < 0.003, FDR = 0.013), glutamine (rho = -0.55; p-value < 0.001, FDR = 0.002), ethanol (rho = 0.64; p-value < 0.001, FDR< 0.001), protein-NH (rho= -0.75; p-value < 0.001, FDR<0.001) among others. HDL-C decreased while IDL-C and VLDL-C increased across all the AP metabolic phenotypes compared to the HC.

**Conclusion:** Dysregulated metabolites and lipids can potentially add to the understanding of the pathophysiological conditions of AP and can aid in the early prognosis and stratification of patients for specialist care.

## 1) Introduction

Acute Pancreatitis (AP) is an inflammatory condition resulting from the autodigestion of the pancreas caused by the sudden release of digestive enzymes before they reach the small intestine. AP clinically presents with nausea, vomiting, abdominal pain, and an increase in amylase and/or lipase in the blood of patients, which is at least three times greater than the normal range (Hofmeyr et al., 2014; Nadhem and Salh, 2017). Globally, the incidence of AP ranges from 5 and 80 people per 100,000 population (Nesvaderani et al., 2015). The incidence of AP in the Western world is well documented, except for Asia and Latin America (Iannuzzi et al., 2022), and low- and middle-income countries such as South Africa, which have a paucity of information (Anderson et al., 2008). Discovery studies from our setting have shown changes in immunological molecules or lack thereof in various stages of the disease (Kay et al., 2017; Thomson et al., 2018, 2019; Nalisa et al., 2021), but incidence studies are still lacking. While there is currently no incidence study of AP in South Africa, hospital prevalence studies have shown that there are increasing hospital admissions due to AP (Funnell et al., 1993; John et al., 1997; Anderson et al., 2008). The most common aetiologies of AP worldwide are biliary and alcohol induced (Zilio et al., 2019). Previously, alcohol-induced AP accounted for most hospital admissions for pancreatitis while antiretroviral drugs (ARVs), endoscopic retrograde cholangiopancreatography (ERCP), hypertriglyceridemia (HTG), or unknown causes together contributed to 5% of the causes of this disease in South Africa (John et al., 1997; Anderson et al., 2008). In 2021, Nalisa and colleagues demonstrated a changing demography of AP aetiology with increasing biliary-related disease which was proportional with alcohol-induced AP (Nalisa et al., 2021). Risk factors such as obesity, age, genetic predisposition, and alcohol abuse differ according to the lifestyle of the population (Yadav and Lowenfels, 2013; Nesvaderani et al., 2015) and play a big role in these changes. In China for example, AP caused by HTG has surpassed alcohol-induced AP due to dietary habits (Yang and McNabb-Baltar, 2020) and it will not be surprising if similar trends are reported in African populations as more Western dietary habits are adopted. Black males have been reported to have a very high risk of AP attributed to social factors such as excessive alcohol consumption (Yang, 2008).

According to the Revised Atlanta Classification (RAC) of 2021, AP is categorized into three classes namely mild AP (MAP), moderately severe AP (MSAP) and severe AP (SAP) (Banks et al., 2013; Choi et al., 2017). The most common form is MAP, which is a self-limiting disease, resolves within the first week or with minimum adverse effect. Moderately severe AP is characterized by organ failure that is transient and resolves within 48 hours (Banks et al., 2013). MSAP patients may develop local complications such as walled-off necrosis, pseudocysts, pancreatic and peripancreatic necrosis, and pancreatic fluid collections (Afghani, 2014). Patients are diagnosed with SAP if organ failure persists for more than 48 hours after admission and/or associated with pancreatic necrosis (Szatmary et al., 2022). Patients with AP are at an increased risk of systemic inflammatory response syndrome (SIRS), due to the activation of the immune system leading to complications such as organ failure and death (Bruen et al., 2021). These patients present with local complications which persist due to the compromised immune system and the resulting immune cascade increases the risk of mortality by 30% (Banks et al., 2013; Yadav and Lowenfels, 2013). Therefore, the severity of the inflammatory response in AP determines which patients will suffer organ failure and thus are classified as severe cases or have a short self-limiting uncomplicated course, classified as MAP.

AP has diverse presentations with clinical variations and an unpredictable course which makes diagnosis and hence prognosis difficult due to changes in the threshold of blood enzymes such as amylase and lipase (Treacy et al., 2001). The varying degrees of severity make it necessary for patients to be evaluated on time in the emergency area for signs and symptoms which will aid in the prevention of organ failure and/or death in patients during hospitalization. This is suggestive of a tight therapeutic window, requiring swift responses, which involves monitoring, intervention and supportive care which is ideally administered within the first 48-72 h from disease onset (Gomatos et al., 2014). Although several rapid, cheap, and reliable tools have been designed to assist clinicians to triage AP patients, sensitivity and specificity challenges associated with the threshold change challenges with blood enzymes means prediction of the disease severity and progression has not been easy and quick enough (Treacy et al., 2001; Magrini et al., 2014; Gu and Tong, 2019; Silva-Vaz et al., 2020). Furthermore, the use of computed tomography (CT) scan may be necessary in cases with diagnostic dilemma and/or where determining the degree of necrosis, which is best demonstrable with imaging obtained after 72 hours from the onset of AP is required (Leppäniemi et al., 2019). The use of these techniques for the diagnosis of critically ill AP patients might delay treatment, which may result in single or multiple organ failure and death, hence the need for more reliable and fast alternatives.

Although, no effective medication exists for the treatment of AP, supportive care which is dependent on the severity is applied (Banks et al., 2006). MSAP and SAP patients who require specialized care are transferred to the intensive care unit (ICU). Antibiotics are administered to patients with infected necrosis (Goodchild et al., 2019). In biliary-associated pancreatitis, surgical interventions such as ERCP with early sphincterotomy can decrease the length of hospital stay and complications (Quinlan, 2014; Chandrasekhara et al., 2017).

The initiation of AP causes alterations in biological processes due to the immune system being activated in response to pancreatic injury (Zheng et al., 2013; Thomson et al., 2018; Nalisa et al., 2021). The activated immune system affects metabolic processes by altering the blood concentration of small molecules such as sugars, lipids, and amino acids (Gu and Tong, 2019; Cacciatore et al., 2021; Elebo et al., 2021). The pancreas secretes digestive enzymes and hormones, which play a key role in the regulation of metabolism, hence damage of pancreatic cells as a result of AP affects the metabolome (Gu and Tong, 2019; Karpińska and Czauderna, 2022). This suggests that metabolomics of biofluids such as plasma and serum may be profiled to retrieve information on the “health” status of the pancreas and pancreas-related diseases aiding in the early detection, diagnosis and identification of novel biomarkers in pancreatic diseases (Gu and Tong, 2019; Siddiqui et al., 2020). The earliest studies on the use of metabolomics to determine metabolites in pancreatitis was the study by Bohus and colleagues and subsequently various other studies have been conducted using different analytical techniques (Bohus et al., 2008; Khan et al., 2013; Xu et al., 2016; Huang et al., 2019, 2022).

Nuclear magnetic resonance (NMR) spectroscopy is a technique used in metabolomics to quantify a wide range of components, including low-molecular-weight metabolites, lipids, and lipoproteins (Sakai et al., 2012; Cacciatore and Loda, 2015; Silva et al., 2020; Elebo et al., 2021). The ^1^H-NMR spectra allows resonances to be assigned directly to the metabolites based on their chemical shifts, and signal multiplicities (Lindon and Nicholson, 2008). Several NMR studies have shown that the integration of metabolites with meta-analysis of the transcriptome revealed a pancreatitis background in pancreatic cancer, congruent with AP being a risk factor to pancreatic cancer (Ketavarapu et al., 2022). Furthermore, ^1^H-NMR metabolomics has been used to distinguish between AP patients and healthy controls or those presenting with abdominal pain (Lusczek et al., 2013; Villaseñor et al., 2014; Shah et al., 2023). In the study by Villaseñor and colleagues in 2014, the authors found changes in the lipid profile with high levels of low-density lipoprotein and unsaturated lipids, which are consistent with known disruptions in lipid metabolism in AP. Most of these studies were carried out in other races and population groups using urine and plasma samples and although the studies targeted metabolites in AP, they did not associate them with severity and outcome. It is also notable that genetic and environmental variations in immune and metabolic responses based on ethnicity means such studies are important in our patient cohort(Yadav and Lowenfels, 2013; Nesvaderani et al., 2015). Here, the serum metabolome of AP patients was profiled for the first time in an African cohort using ^1^H-NMR spectroscopy. This study aimed to investigate altered metabolites and lipids to understand the underlying pathophysiological conditions of AP in the three AP severities to enable improved prognosis, for early patient stratification and management.

## 2 Materials and methods

### 2.1 Sample collection and processing

Ethics clearance for this study was obtained from the Human Research Ethics Committee (Medical) of the University of the Witwatersrand (Ethics No. M190407). The RAC of 2012 (MAP, MSAP, or SAP) was used by consulting clinicians to recruit participants at the Chris Hani Baragwanath Academic Hospital (CHBAH). To be included in the study, participants had to be 18 or more years old and self-identified as of African descent. Patients were excluded if they had been diagnosed with auto-immune diseases, undergoing immunotherapy, and were on antibiotics within one month of admission. Following informed consent from the participants, blood samples were collected using red coloured non-anticoagulant BD vacutainer® blood collection tubes (BD Biosciences, New Jersey, USA). Four millilitres (4mL) of blood was drawn from eligible patients as previously described by Nalisa and colleagues on days 1, 3, 5, and 7 post epigastric pain enabling time points and longitudinal analysis of the metabolic changes (Nalisa et al., 2021). Patients typically reported to hospital on Day 3 of pain.

The clinical data were obtained from the patient’s hospital files, via the National Health Laboratory Services (NHLS) Trakcare Web Results, version L6.10, captured on the RedCap® (Tennessee, USA) database. Captured data included demographic, body mass index (BMI), AP aetiology, AP severity, clinical test results, and comorbidities. Healthy volunteers were recruited as controls and were age and sex-matched with the AP patients. The collected blood samples were allowed to stand upright for 30 minutes at room temperature in blood collection tubes and then centrifuged at 1734 *xg* at 4°C for 30 minutes. The serum was aliquoted into microfuge tubes of 500 µL volumes and stored at -80°C until needed for the analysis.

### 2.2 Serum sample preparation

The frozen serum samples, previously processed and stored at -80°C were left to thaw at room temperature. A working solution was prepared by adding 300 µL of the thawed serum to 300 µL of a solution consisting of 0.75 M potassium phosphate buffer (pH 7.4), 5.81 mM of trimethylsilyl-2,2,3,3-tetradeuteropropionic acid (TSP; Sigma-Aldrich, St. Louis, MO, USA) and a trace amount of sodium azide (65 mg dissolved in deuterium oxide) to prevent bacterial growth. The samples were vortexed to obtain a homogenous mixture. A final volume of 540 µL of each sample was transferred to a 5 mm NMR tube (Wilmad Lab Glass, Vineland, NJ, USA) for analysis.

### 2.3 Lipid Extract Preparation

The extraction of the lipids was performed using the protocol described by Lamiquiz-Moneo and colleagues (Lamiquiz-Moneo et al., 2019). A volume of 300 µL of BUME (butanol: methanol—2:1) was added to 100 μL serum aliquots in glass GC vials as well as 300 μL DIPE (diisopropyl ether: ethyl acetate—3:1) and 300 μL HCO. Firstly, BUME and HCO were added to the samples and vortexed for 1 minute. DIPE was then added to extract the lipids and the mixture incubated on a shaker for 10 minutes. To separate the lipids from the mixture, the samples were centrifuged at 2058 xg for 5 minutes. Afterwards, the top layer was transferred to clean vials, dried under liquid nitrogen at 37 °C and then resuspended in 600 μL solution of CDCl3:CD3OD:DCO (chloroform-d:methanol-d:water-d; 16:7:1, v/v/v) containing 1.18 mM TSP. A volume of 540 μL of lipid extract samples were transferred to 5 mm NMR tubes (Wilmad Lab Glass) and analysed using NMR spectroscopy.

### 2.4 NMR Analysis

The NMR tubes containing the diluted metabolite and lipid extracts, respectively were loaded on a 500 MHz Bruker Avance III HD NMR spectrometer equipped with a triple-resonance inverse 1H probe head and x,y,z gradient coils. Afterwards, one-dimensional (1D) proton (1H)-NMR spectra were acquired using different pulse sequences. A standard nuclear overhauser effect spectroscopy (NOESY) pulse sequence presat (noesygppr1d.comp) was used on both the metabolite and lipid extract samples. NOESY was used to detect both signals of small metabolites and high-molecular-weight macromolecules such as lipoproteins. Additionally, a standard diffusion-edited (DIFF) pulse sequence (ledbpgppr2s1d) was used to detect only high-molecular-weight macromolecules, such as lipoproteins. Pooled AP samples of different severities were used as a quality control sample and were included in each batch for qualitative assessment of repeatability by overlaying the raw spectra.

### 2.5 NMR Profiling

NMR spectroscopy was used to quantify a panel of 38 signals. The peaks of the identified metabolites were fitted by combining a local baseline and Voigt functions based on the multiplicity of the NMR signal (Marshall et al., 1997). The assignment of quantified signals is reported in **Table S1**. To validate the efficacy of the different deconvolution models, the root-mean-square deviation was determined. The absolute concentration of each metabolite was calculated according to a previously reported equation (Serkova et al., 2005). The number of protons contributing to the unknown signals was imputed to 1. The concentration of carbohydrates was also estimated by considering the equilibrium between their cyclic forms.

N-acetylglucosamine/galactosamine (GlycA) and N-acetylneuraminic acid (GlycB) signals were quantified as inflammatory markers by integrating the areas between 2.00 and 2.05 ppm and 2.09 and 2.05 ppm, respectively. GlycA is measured as an NMR signal of post-translational modification of glycosylated acute-phase proteins released during inflammation (Otvos et al., 2015). The Liposcale test (Biosfer TesLab, Reus, Spain) was then used to determine lipoprotein parameters, high-density lipoprotein (HDL), low-density lipoprotein (LDL), and very low-density lipoprotein (VLDL) particle number, size, and lipid concentration of each subtype (Mallol et al., 2015), was used as previously described by (Elebo et al., 2021). Each of the DIFF spectrum in the range between 0.1 and 9.5 ppm, excluding the regions corresponding to the water signals between 4.40 and 5.00 ppm, was segmented into 0.001-ppm chemical shift bins, and the corresponding spectral areas under the curve, giving a total of 8800 variables.

### 2.6 Statistics and Data Analysis

Statistical analysis and graphical illustrations of the data were generated in R (version 3.6.1) and R studio (version 1.1.456) software using scripts developed in-house. Wilcoxon and Kruskal-Wallis rank-sum tests were used to compare differences in numerical covariates (e.g., age and metabolite concentration). Fisher’s exact test was used to assess differences between categorical variables (e.g., gender), and Spearman’s rank test was then used to calculate the correlation coefficient (rho) between variables. P-values < 0.05 were considered significant and to account for multiple testing, a false discovery rate (FDR) of <10% was applied.

The KODAMA algorithm, which allows for unsupervised extraction of features and enables analysis of noisy datasets of high dimension was used to facilitate the identification of patterns representing underlying metabolic phenotypes on all samples (Cacciatore et al., 2014, 2017; Bray et al., 2017). The partition around medoids (PAM) clustering (Reynolds et al., 2006) was applied to the KODAMA scores using the silhouette algorithm 10 (Rousseeuw, 1987) to verify the results obtained. The silhouette median value was utilized to assess the ideal number of clusters, ranging from 2 to 10.

Using partial least-squares (PLS) analysis, regression was performed on DIFF spectra metabolic profiles. Furthermore, 10-fold cross-validation was performed to evaluate the predictive efficacy of the model (Bertini et al., 2012). Both the goodness of fit parameter (R^2^) and the predictive ability parameter (Q^2^) were also calculated using standard formulas (Eriksson et al., 2003). The Q^2^ value was calculated from the p-value to assess the performance of the PLS regression model (Szymańska et al., 2012). A p-value < 0.05 was regarded as significant.

## 3 Results

### 3.1 Demographics and Clinical Characteristics

#### 3.1.1 Patients’ demographic and clinicopathological characteristics

Thirty (30) patients with AP (MAP = 8, MSAP = 14, and SAP = 8) met the inclusion criteria and 9 healthy controls (HC) were all enrolled in the study. There were equal number of males (11) and females (11). There were no statistically significant differences in BMI, age, and gender across the four groups as shown in **Table 1**. The primary aetiology of AP in this study was alcohol-induced (16/30 = 53%) occurring mostly in males (12/16=75%). Biliary associated-AP was the second common aetiology (12/30 = 40%) with majority being female (9/12=75%), and ARV-induced AP was 7% of the population (2/30) as shown in **Table S2**. In this study, 50% of AP patients with ARV-associated aetiology were women.

**Table 1:** Demographic and clinical characteristics of the acute pancreatitis patients.

| Feature | Control (n=9) | MAP (N=8) | MSAP (N=14) | SAP (N=8) | p-value |
| --- | --- | --- | --- | --- | --- |
| BMI (kg/m<sup>2</sup>), median [IQR] | 30.4 [23.6<br>32.8] | 24.8 [24.7 44.3] | 27 [23.2 30.5] | 32 [30.9 39.8] | 0.515 |
| Age (years), median [IQR] | 43 [30 54] | 43.5 [36.5 54.8] | 38 [32 43.5] | 48 [32.2 57.2] | 0.613 |
| Gender |  |  |  |  | 0.100 |
| F, n (%) | 5 (55.6) | 4 (50.0) | 7 (50.0) | 4 (50.0) |  |
| M, n (%) | 4 (44.4) | 4 (50.0) | 7 (50.0) | 4 (50.0) |  |
| Aetiology of pancreatitis |  |  |  |  |  |
| Biliary, n (%) |  | 4 (50.0) | 5 (35.7) | 3 (37.5) |  |
| Alcohol, n (%) |  | 4 (50.0) | 8 (57.1) | 4 (50.0) |  |
| ARVs, n (%) |  | 0 (0.0) | 1 (7.1) | 1 (12.5) |  |
| Days of hospitalization, median [IQR] |  | 10.5 [4.8 15] | 9 [4.8 11] | 8 [8 10] | 0.864 |
| no, n (%) |  | 7 (87.5) | 10 (71.4) | 8 (100.0) |  |
| yes, n (%) |  | 1 (12.5) | 4 (28.6) | 0 (0.0) |  |
| Diabetic |  |  |  |  | 0.100 |
| no, n (%) |  | 8 (100.0) | 13 (92.9) | 8 (100.0) |  |
| yes, n (%) |  | 0 (0.0) | 1 (7.1) | 0 (0.0) |  |
| HIV |  |  |  |  | 0.650 |
| no, n (%) |  | 7 (87.5) | 10 (71.4) | 7 (87.5) |  |
| yes, n (%) |  | 1 (12.5) | 4 (28.6) | 1 (12.5) |  |
| Organ dysfunction |  |  |  |  | 0.027 |
| no, n (%) |  | 8 (100.0) | 8 (57.1) | 3 (37.5) |  |
| renal, n (%) |  | 0 (0.0) | 0 (0.0) | 1 (12.5) |  |
| respiratory, n (%) |  | 0 (0.0) | 3 (21.4) | 4 (50.0) |  |
| transient renal, n (%) |  | 0 (0.0) | 3 (21.4) | 0 (0.0) |  |
| Local Complications |  |  |  |  | 0.878 |
| ANP, n (%) |  | 1 (12.5) | 0 (0.0) | 0 (0.0) |  |
| no, n (%) |  | 7 (87.5) | 10 (76.9) | 6 (85.7) |  |
| pancreatic collection, n (%) |  | 0 (0.0) | 1 (7.7) | 0 (0.0) |  |
| peripancreatic fatty stranding, n (%) |  | 0 (0.0) | 0 (0.0) | 1 (14.3) |  |
| wall off necrosis; ANC, n (%) |  | 0 (0.0) | 1 (7.7) | 0 (0.0) |  |
| yes, but not reported, n (%) |  | 0 (0.0) | 1 (7.7) | 0 (0.0) |  |
| Admission to ICU |  |  |  |  | 0.004 |
| no, n (%) |  | 8 (100.0) | 13 (92.9) | 3 (37.5) |  |
| yes, n (%) |  | 0 (0.0) | 1 (7.1) | 5 (62.5) |  |
| Surgical procedures |  |  |  |  | 0.566 |
| cholecystectomy, n (%) |  | 2 (28.6) | 1 (7.7) | 0 (0.0) |  |
| ERCP, n (%) |  | 0 (0.0) | 1 (7.7) | 1 (14.3) |  |
| None, n (%) |  | 5 (71.4) | 11 (84.6) | 6 (85.7) |  |
| Hospital death |  |  |  |  | 0.02 |
| no, n (%) |  | 8 (100.0) | 13 (92.9) | 4 (50.0) |  |
| yes, n (%) |  | 0 (0.0) | 1 (7.1) | 4 (50.0) |  |

**Abbreviations.** ARVs- antiretrovirals, HIV- human immunodeficiency virus, ANP- Atrial natriuretic peptide, ANC- absolute neutrophil count, ICU- intensive care unit, ERCP Endoscopic retrograde cholangiopancreatography, IQR- The Interquartile Range.

The incidence of organ dysfunction (OD) increased with the severity of AP, with 50% of MSAP patients having respiratory OD as shown in **Table 1**. Imaging findings consistent with local complications, such as pancreatic collection and wall-off necrosis, were found in some MSAP and SAP patients. As expected, admissions to the Intensive Care Unit (ICU) and hospital death were prevalent in SAP patients.

#### 3.1.2 Routine clinical / Biochemical tests

No statistically significant correlation was observed between the severity of AP and routine biochemical tests such as amylase, lipase, CRP, haemoglobin, haematocrit and platelets (**Table S3**). The median values of liver function parameters; total bilirubin, conjugated bilirubin, alanine transaminase (ALT), aspartate transaminase (AST), and alkaline phosphatase (ALP) were observed to be elevated above the physiological range for SAP patients. A third (10/30) of the patients in our cohort were affected by hypoalbuminemia, in contrast, no correlation with severity was reported with CRP (**Figure 1A**). In this cohort, amylase and ALP (p<0.001) showed statistically significantly higher concentrations in biliary-associated AP with their combination able to differentiate biliary-associated from alcohol-associated AP as demonstrated in **Figure 1B**.

**Figure 1.**
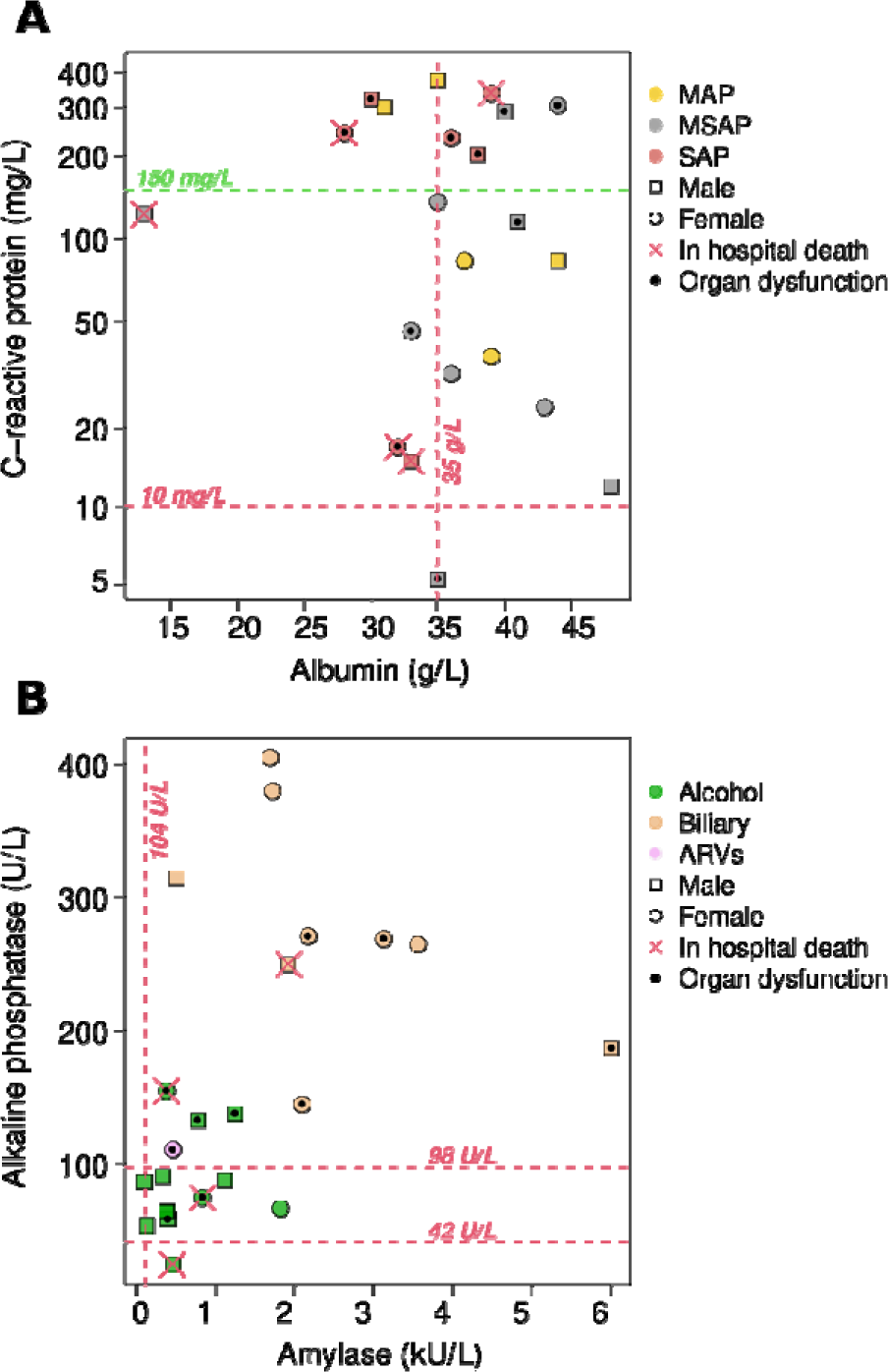
Plot showing (**A**) correlation between albumin and CRP: The top and middle right quadrants are patients with Glasgow Prognostic Score (GPS) =1 with CRP> 150mg/L and albumin levels > 35g/L while the left quadrants are patients with GPS =2. Although the top right quadrant has GPS=1, the patients are characterised with an elevated organ dysfunction levels and (**B**) the correlation between amylase and ALP: Decreased levels of both amylases and alkaline phosphatase (ALP) is associated with alcohol-associated AP while the increased levels are linked to biliary-associated AP. GPS of 0, 1 or 2 refers to neither, one or both abnormalities.

Most AP patients in this cohort showed a very high values of CRP (>10 mg/L). AP patients with CRP higher than 150 mg/L and albumin lower than 35 g/L were more likely to experience in-hospital death or have organ dysfunction (11/13=85%). A comparison of the biochemical tests showed that both amylase (*p*-value <0.001; FDR= 0.008) and ALP (*p*-value <0.001; FDR= 0.003) were statistically different among the 3 different aetiologies of AP (**Table S4**). The Glasgow Prognostic Score (GPS) reflects the systemic inflammatory response, derived from the serum concentrations of CRP, and nutritional status, derived from the albumin levels (Forrest et al., 2003). This score ranges from 0 to 2 with a higher value implying the presence of both abnormalities high CRP and low albumin levels. Based on our reported CRP and albumin levels, 32% (7/22) of AP patients had a GPS of 2, while 64% (14/22) AP patients had a GPS score of 1 and 4% of patients had a GPS of 0. Furthermore, there were no statistically significant differences observed for a comparative analysis between the different AP severity groups and biochemical tests performed in biliary-associated AP (**Table S5**) and alcohol-associated AP (**Table S6**).

### 3.2 Metabolites and acute pancreatitis severity

NMR spectroscopy was used to profile the concentration of metabolites and lipids and in total 38 metabolic signals, including lipid classes and the NMR inflammatory markers, GlycA and GlycB, were reported. To delineate the metabolic signatures of AP, the Spearman correlation test was performed to link metabolic parameters to the different groups in the following rank order: HC=0, MAP =1, MSAP =2, and SAP = 3 (**Table 2)**. Elevated levels of 3-hydroxybutyrate (3-HB), acetoacetate, phenylalanine, mannose, and lactate, as well as decreased concentrations of ascorbate, methanol, ethanol, and glutamine was observed with increased AP severity. Additionally, AP severity showed a large alteration in lipid profile with increases lipid alpha-CH2 and decreases in cholesterol, Lipid=CH-CH2-CH=, Lipid beta-CH2 and lipid CH3 respectively with diseases severity as well as a depletion of the serum levels of protein (Protein NH). Furthermore, three unknown signals; 1.45, 1.11 and 1.06 ppm were observed to be statistically significant. Although not statistically significant, inflammatory markers GlycA and GlycB showed a positive correlation with the severity of the disease. The aetiology of AP, i.e., alcohol or biliary-induced, did not have a significant impact on the metabolic profile (**Table S7**).

**Table 2.** Correlation of concentrations of metabolites with the severity of acute pancreatitis.

| Feature | rho | p-value | FDR |
| --- | --- | --- | --- |
| Formate | 0.04 | 0.793 | 0.809 |
| Unknown signal at 8.12 ppm | 0.01 | 0.951 | 0.951 |
| Unknown signal at 8.07 ppm | -0.08 | 0.630 | 0.684 |
| **Phenylalanine** | **0.54** | **<0.001** | **0.028** |
| Tyrosine | -0.13 | 0.443 | 0.525 |
| Unknown signal at 7.14 ppm | 0.19 | 0.237 | 0.318 |
| Histidine | -0.1 | 0.557 | 0.631 |
| Glucose | 0.24 | 0.140 | 0.238 |
| **Mannose** | **0.57** | **<0.001** | **0.015** |
| Unknown signal at 5.15 ppm | 0.23 | 0.155 | 0.239 |
| Unknown signal at 5.09 ppm | 0.16 | 0.330 | 0.421 |
| Unknown signal at 5.01 ppm | 0.23 | 0.155 | 0.239 |
| **Ascorbate** | **-0.46** | **0.003** | **0.013** |
| Threonine | -0.33 | 0.043 | 0.104 |
| **Lactate** | **0.67** | **<0.001** | **<0.001** |
| Creatinine | 0.2 | 0.218 | 0.309 |
| Creatine | 0.3 | 0.064 | 0.143 |
| Glycine | 0.06 | 0.717 | 0.746 |
| **Methanol** | **-0.46** | **0.001** | **0.013** |
| Unknown signal at 2.55 ppm | 0.25 | 0.130 | 0.229 |
| Citrate | -0.16 | 0.330 | 0.421 |
| **Glutamine** | **-0.55** | **<0.001** | **0.003** |
| Pyruvate | 0.2 | 0.230 | 0.317 |
| Glutamate | 0.25 | 0.126 | 0.229 |
| **Acetoacetate** | **0.63** | **<0.001** | **<0.001** |
| Acetate | 0.28 | 0.089 | 0.169 |
| Alanine | -0.29 | 0.077 | 0.158 |
| Unknown signal at 1.45 ppm | 0.44 | 0.005 | 0.016 |
| Unknown signal at 1.43 ppm | 0.23 | 0.155 | 0.239 |
| **3-Hydroxybutyrate** | **0.46** | **<0.001** | **0.013** |
| **Ethanol** | **-0.64** | **<0.001** | **<0.001** |
| Unknown signal at 1.16 ppm | -0.07 | 0.686 | 0.729 |
| Unknown signal at 1.14 ppm | 0.31 | 0.054 | 0.124 |
| Unknown signal at 1.11 ppm | 0.47 | 0.003 | 0.013 |
| Unknown signal at 1.06 ppm | 0.4 | 0.011 | 0.032 |
| Valine | -0.14 | 0.411 | 0.511 |
| Isoleucine | 0.22 | 0.018 | 0.276 |
| Leucine | 0.11 | 0.049 | 0.564 |
| 2-Hydroxybutyrate | 0.34 | 0.032 | 0.081 |
| **Protein NH** | **-0.75** | **<0.001** | **<0.001** |
| Unsaturated lipid -CH=CH- | -0.35 | 0.029 | 0.077 |
| **Lipid alpha-CH2** | **0.45** | **0.004** | **0.013** |
| **Cholesterol** | **-0.43** | **0.006** | **0.017** |
| **Lipid =CH-CH2-CH=** | **-0.55** | **<0.001** | **0.002** |
| Glycerol phospholipid | 0.21 | 0.199 | 0.289 |
| Phospholipid | -0.13 | 0.440 | 0.525 |
| **Lipid beta-CH2** | **-0.52** | **<0.001** | **0.004** |
| Lipid CH2 | 0.09 | 0.599 | 0.664 |
| **Lipid CH3** | **-0.44** | **0.005** | **0.016** |
| GlycB | 0.28 | 0.085 | 0.167 |
| GlycA | 0.29 | 0.074 | 0.158 |

Rho: Spearman's correlation coefficient; FDR: False Discovery rate

### 3.3 The Relationship between the metabolic phenotype and clinical outcomes of acute pancreatitis

Unsupervised KODAMA analysis was performed to visualize patterns between metabolic phenotypes (metabotype) and disease severity as described in **Figure 2A**. This operation separated the patient cohort into three different metabotypes. A distinct metabolic profile was observed for the healthy controls compared to AP patients. Notably, a clear separation was also evident between patients with MAP and SAP. Interestingly, two patients with SAP showed a metabolic profile similar to the MAP. Further analysis indicated that these patients had elevated levels of free cholesterol/cholesterol ester ratio, which could be due to cholestasis linked to lipoprotein X (LpX). Cholestasis is characterized by the presence of Lpx (Brandt et al., 2015; Phatlhane and Zemlin, 2016), that could mask the SAP phenotype. The group of AP patients with intermediate severity (i.e., MSAP) shared metabolic profiles with both MAP and SAP patients (**Figure 2B**). However, MSAP patients with a closer phenotype to SAP had a higher incidence of adverse events (**Figure 2C**). Some of the adverse events were complications such as infection and pseudocysts, organ dysfunction (OD), and admission to the ICU. Notably, increases in lactate activity, shown by the colour changes from blue through yellow in the heatmap (**Figure 2D)**, which is prevalent in the SAP patients also corresponds with increase OD, local complications, ICU admission and death.

**Figure 2:**
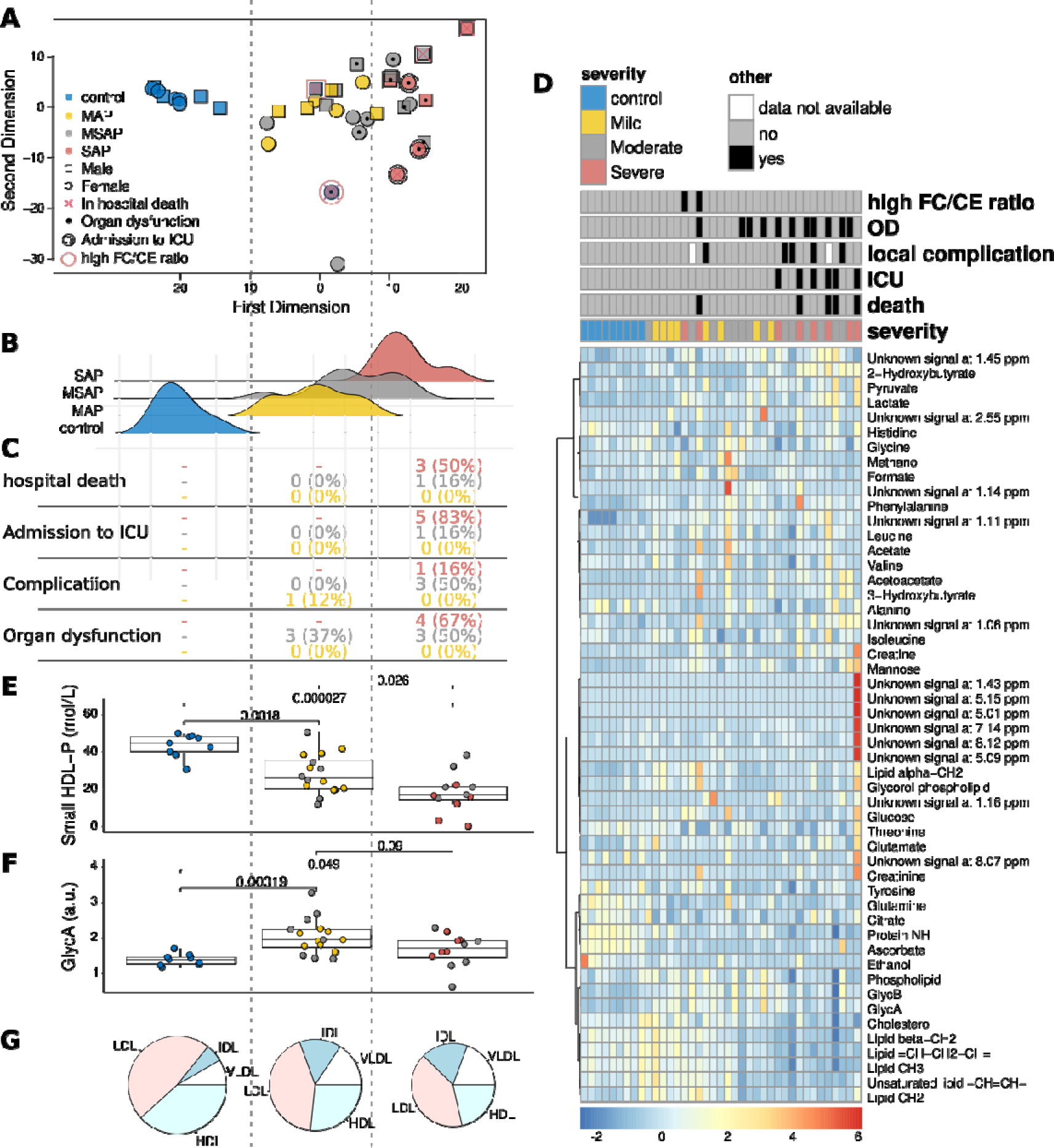
Overview of the metabolic phenotypes of AP and their effects. **A;** Unsupervised analysis of the metabolites of HC (n = 9), MAP (n = 8), MSAP (n = 14), and SAP (n = 8). Two distinct clusters representing the metabolic phenotype of mild and severe AP separated with most of the MAP and about 5 of the MSAP in the first component while most of the SAP except for 2 patients are in the second component (components are delineated throughout relevant figures using broken vertical lines). **B;** The distribution of the different AP and HC groups, the distinct separation of the HC and SAP was observed at the left and right of the graph respectively. Both MAP and MSAP were observed in the middle with MAP at the left end and the MSAP at the right end. **C**; Comparison of the incidence of the clinical features across the groups. MAP had the least adverse effect and complications while SAP presented the most adverse effects including death. MSAP had the highest levels of organ dysfunction and complication in hospitalized patients. **D**; Heatmap showing the comparison of the clinical features and the metabolites across the AP groups. Lipid profiles were observed to distinctly separate the AP groups as colour changes were observed from blue to red. **E** and **F**; small HDL-P and GlycA across the metabolic phenotypes of AP. Decreased levels of small HDL-P with increased AP severity were observed. GlycA increased for the mild pancreatic phenotype and reflects a moderated decline for the severe phenotype. **G**; Distribution of cholesterol related lipids across the various components from HC, MAP/MSAP and MSAP/SAP. HDL-C decreased from HC to SAP while both VLDL-C and IDL-C were elevated. Decreased HDL-C in AP might represent impaired anti-inflammatory function, reduced lipoprotein synthesis, and impaired immune functions. HDL-High-density lipoprotein, HDL-P-HDL-Particle, LDL-Low-density lipoprotein, VLDL-Very-Low Density Lipoprotein, IDL – immediate-density lipoprotein.

A further comparison of the clinical features and the metabolite profile showed that the lipid profile distinctively separated the metabolic phenotypes of AP based on their severity with an overall decrease in lipid levels with increasing AP severity (**Figure 2D**). The concentration of high-density lipoprotein particles (HDL-P) seemed to constitute the difference in the dyslipidaemia found between the metabolic phenotypes, with a notable decrease with increasing AP severity (**Figure 2E**). The inflammatory marker, GlycA, increased across the group and was highest in the MSAP patients (**Figure 2F**). After further characterisation of cholesterol-related lipids, it was shown that dysregulation of HDL (decreasing with severity) and IDL and VLDL (both increasing with severity), were linked to AP severity (**Figure 2G**).

### 3.4 Trend Analysis

To further understand the association of metabolites and lipoproteins with AP severity, KODAMA analysis was used to analyse three representative molecules (GlycA, 3-hydroxybutyrate and lipid alpha-CH2). A comparison of the metabolites (**Table S8**) and lipoproteins (**Table S9**) with inverted first dimension KODAMA showed dysregulated metabolites such as mannose, ascorbate, phenylalanine and lipoproteins among others as we move from MAP to SAP. Some of these metabolites were also dysregulated over time as days progressed from days 1 – 7 for selected severities albeit inconsistently for example 3-hydroxybutyrate in SAP patients. The correlation of the concentration of metabolites with AP severity showed that GlycA (p-value = 0.004) for example, was significantly altered (**Figure 3A**, **Table S10**). In this pilot study, the ketoacidosis metabolite, 3-hydroxybutyrate (p-value = 0.003) was shown to be significantly linked to worse clinical outcomes in AP as shown in **Figure 3B** (**Table S11**) and within the severe group this seems to increase with time from day1 to day 7. Elevated levels of 3-hydroxybutyrate were observed in SAP patients compared to the other severity groups. A metabolite of interest, Lipid alpha-CH2 was observed to be dysregulated in AP patients with organ dysfunction (**Figure 3C**). Elevated levels of alpha-CH2 (p-value = 0.007) were seen in SAP compared to MSAP and MAP (**Table S12**). Within patients, it appeared to be consistent over time.

**Figure 3:**
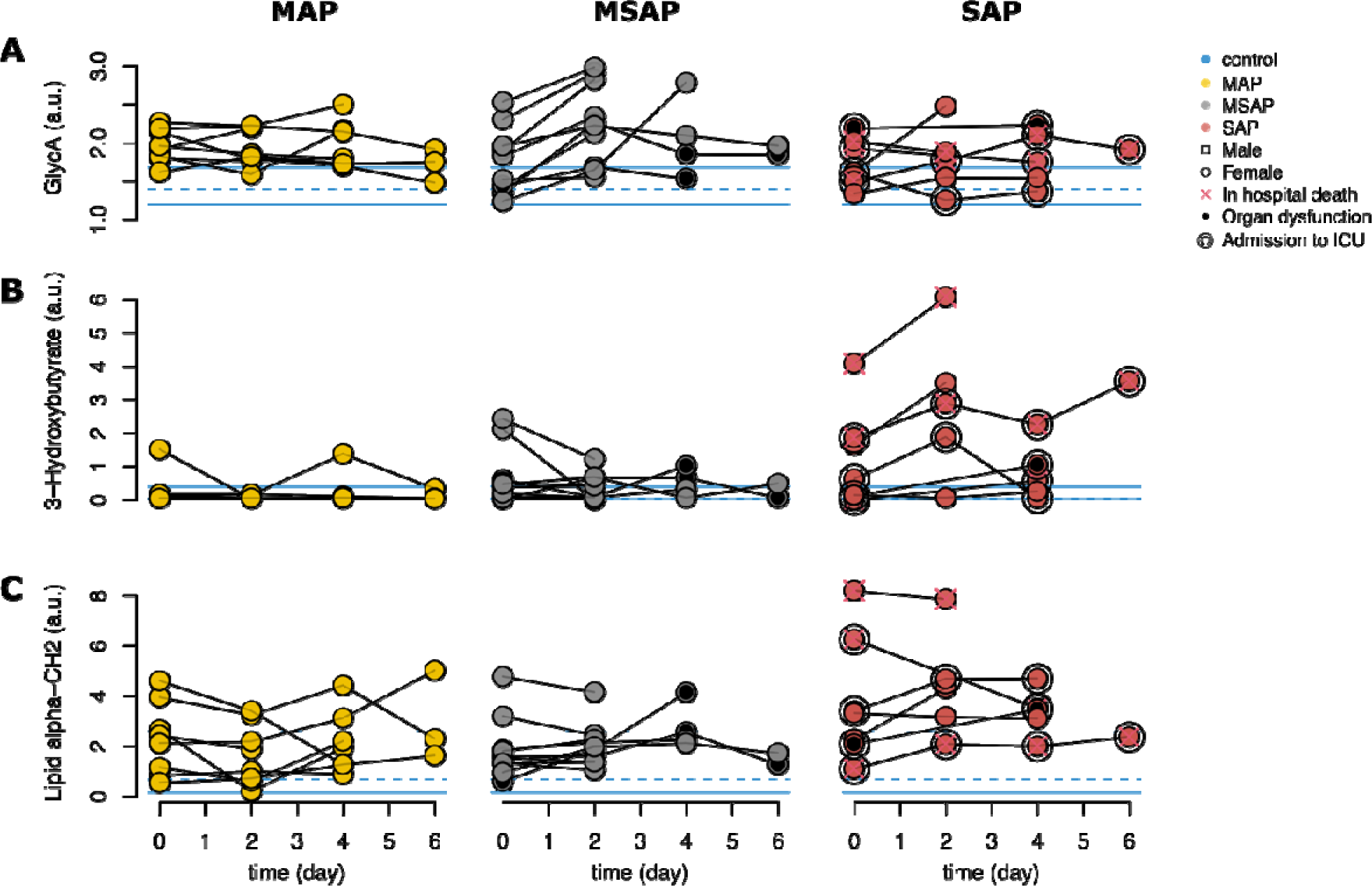
Trends in selected metabolic parameters. **A**; describes the pattern for GlycA in the severity groups of AP over all time points. **B**; assess levels of 3-hydroxybutyrate in the severity groups of AP over all time points. **C**; describes the pattern for lipid alpha-CH2 in the severity groups of AP over all time points.

## 4 Discussion

In this preliminary study, the demographics, clinicopathological characteristics, as well as the metabolite and lipoprotein profiles of 30 AP patients were analysed to assess the relationship of these parameters to AP severity and clinical outcome. Importantly, the identification of a metabotype that could help in prognosticating AP disease severity was envisaged.

Approximately 57% (17/30) of the AP patients were obese with a BMI of 32 kg/m ² and a median IQR of 30.9 - 39.8 kg/m ² in SAP. Notably, obesity was more common in patients with biliary-associated AP. An increase in BMI of about 83% (10/12) was observed for the biliary-associated AP. Previous studies have shown that comorbidities such as obesity affect the clinical outcomes of AP (Khatua et al., 2017; Moran et al., 2018), and this is also true for the cohort evaluated.

Of interest, this study showed that alcohol-induced AP is the main aetiology of male AP patients while biliary associated AP is common in females. On the contrary, a retrospective study in a Western society showed biliary-induced AP, which is prevalent in females, to be the main cause of AP followed by alcohol, which was predominantly in males (Nesvaderani et al., 2015). Furthermore, over 50% of the female AP patients in this study were living with HIV/AIDS and on antiretroviral therapy (ART). Acute pancreatitis is extremely high in people living with HIV/AIDS (PLWHA), which could be due to the long-term use of antiretroviral therapy (ART) medications such as corticosteroids and sulphonamides (Dragovic, 2013). Importantly, the aetiology of AP did not have a notable impact on the metabolic profile as shown in Table S7, and this might be because the course of the disease is not usually affected by the aetiology (Fonteh et al., 2018).

Serum amylase and lipase are routine tests used for the diagnosis of AP (Ismail and Bhayana, 2017). According to the South African NHLS report, the physiological concentrations of amylase and lipase range from 28 -110 U/L and 13 - 60 U/L respectively (National Health Laboratory Services 2019/20 Annual Report; National Health Amendment Bill: rejection | PMG, n.d.). Our study participants had higher concentrations of both amylase and lipase, about three times above the normal limit but no statistically significant differences were found between the groups probably because of the small sample sizes. Elevated levels of amylases and ALP were observed in our study population, which suggests liver damage or a blocked bile duct in biliary pancreatitis (Fratantoni et al., 2021) hence these findings could be vital in distinguishing from alcohol-induced AP (Hong et al., 2017). Indeed, this study confirms that the combination of amylase and ALP can be used to differentiate biliary-associated AP from alcohol-associated AP (Güngör et al., 2011; Thiyagarajan and Amirthavarshini Ponnuswamy, 2020).

This study further confirms the previously reported direct association of hypoalbuminemia with severity and mortality in AP (Ocskay et al., 2021). Elevated inflammatory states have been shown to decrease albumin levels via capillary leak or induction of shorter half-life associated with increased severity (Soeters et al., 2019; Ocskay et al., 2021). In contrast, there was no correlation reported between CRP and severity in AP in this cohort. This is not surprising given that there is conflicting evidence in the literature on the role of CRP in discriminating complicated AP early in the disease (Ahmad et al., 2021). Rather, CRP is considered useful in predicting severity when used in combination with other scoring systems to differentiate mild from severe patients for early decision making. Most AP patients in this cohort showed high values of CRP (>10 mg/L). Based on these values, we further analysed the patients for systemic inflammation and nutritional status using the GPS system. The data showed that 32% (7/22) of AP patients were classified with a GPS of 2, 64% (14/22) with a GPS of 1; only 4% of patients were classified with a GPS of 0. From the results, MSAP patients had the most complications and organ failure from pancreatic necrosis, which ultimately results in organ failure if unresolved (Niknam et al., 2020). SAP had the most number of deaths, which is unsurprising given that SAP is characterized by persistent organ failure for over 48 hours (Foster et al., 2016). Most of these SAP patients also generally had higher CRP levels and hypoalbuminemia. The disease progression leading to organ failure and/or death in AP is usually due to SIRS, which is a response to an infection or an injury in the pancreas (Garg and Singh, 2019).

In this study, the physiological metabotype created was able to distinguish between the mild and the severe groups of patients. AP patients undergo a hypermetabolic condition, which dysregulates amino acid, lipid, and glucose metabolism (Kotan et al., 2023). Metabolites such as lactate, phenylalanine, mannose, and acetoacetate, and 3-HB increased in concentration while ascorbate, methanol, ethanol, and glutamine reduced with AP severity in this cohort. Similar findings have been reported by Li *et al*., (2014) and Xiao *et al*., (2017) in various AP settings (Li et al., 2014; Xiao et al., 2017).

Lactate levels were the most positively correlated metabolite with AP severity. Oxygen deficiency and increased energy consumption enhances anaerobic glycolysis, which occurs in the early phase of AP ultimately leading to elevated levels of lactate, a glycolytic metabolite (Li et al., 2014). Increased lactate concentration has been associated with persistent organ failure in AP (Valverde-López et al., 2017) and is independently associated with poor outcomes and death (McDonald et al., 2017). Increases in mannose levels also positively correlated with AP severity and are not surprising given the involvement of the glycolytic pathway (Xiao et al., 2017).

Organ failure and mortality have been associated with raised phenylalanine levels in the plasma of AP patients (Chen et al., 2020; Xu et al., 2020). Increases in phenylalanine levels could be as result of scavenging of tetrahydrobiopterin, a key co-factor involved in the conversion of phenylalanine to tyrosine (Sandstrom et al., 2008). Dysregulated amino acid metabolism leads to the release of excess acetyl CoA into the tricarboxylic acid (TCA) cycle, which is converted to ketone bodies such as 3-HB (Li et al., 2014). Ketones, which are products of the oxidative pathway of alcohol metabolites could be predictive of alcohol-induced AP (McGuire, 2006; Villaseñor et al., 2014). Elevated levels of 3-HB in AP has been shown to differentiate between MAP and SAP (Xiao et al., 2017) and associated with poor survival rates in pancreatic cancer (Elebo et al., 2021). In this study, the 3-HB increased with severity and was found to persist in the SAP patients over time when a trend analysis was performed. Hydroxybutyrate dehydrogenase enzyme, which catalyses the breakdown of 3-HB to acetoacetate, another ketone (Hoque et al., 2009), has been shown to significantly increase with organ failure and SIRS (Xiao et al., 2020). The increasing levels of both 3-HB and acetoacetate with increasing severity is suggestive of the reversible reaction between the two molecules with the possibility of an equilibrium. Hence, both 3-HB and acetoacetate could be used as a clinical biomarker to predict worse outcomes.

It is not surprising that ascorbate (vitamin C) levels decreased in this study with increasing AP severity, a phenomenon that has been previously reported (Gao et al., 2021). Due to this deficiency in ascorbate in AP, high doses of ascorbate have shown therapeutic efficacy in AP (Du et al., 2003). Glutamine levels also significantly decreased with increasing AP severity. The consumption of glutamine by immune cells is similar if not more than that of glucose and it is essential for processes such as the proliferation of immune cells and the production of cytokines amongst others (Cruzat et al., 2018). (The inflammatory and immune compromised patient, therefore, inevitably experiences decreasing levels with severity as was the case here.

The level of two alcohols, methanol and ethanol, decreased with increasing AP severity but just like the rest of the metabolites, were not significantly different between biliary and alcohol induced AP patients (**Table S7**) as would have been expected. This lack of a strong metabolic signature is supported by the fact that irrespective of the cause of AP, the trajectory is usually the same (Fonteh et al., 2018), hence aetiology does not influence metabotype. In addition, the mechanisms underlying the aetiology of AP have not been extensively elucidated especially for alcohol-induced AP and could potentially shed light on this. This study showed a dyslipidaemic profile with AP groups distinctly based on their severity. Serum lipid levels are associated with the severity of acute pancreatitis (Khan et al., 2013; Zhang et al., 2017). Small HDL-P decreased with increased severity of AP. Studies have shown that small HDL-P played a vital role in the ATP binding Cassette transporter A 1 (ABCA-1) mediated cholesterol efflux from macrophages, which leads to the deployment of intracellular cholesterol to the plasma membrane (Du et al., 2015). HDL-C decreased from HC to SAP while IDL and VLDL increased with severity. Recent studies demonstrated that decreased HDL-C levels are an independent prognostic factor of adverse outcomes such as persistent organ failure and death (Zhang et al., 2017). HDL-C possesses anti-inflammatory properties, hence decreased amounts could be latent for AP severity (Murphy and Woollard, 2010). Oxidized VLDL is a marker of oxidative stress which inhibits lipoprotein lipase responsible for triglyceride (TG) metabolism (Jong et al., 2000). Elevated VLDL levels are associated with HTG which is the third leading cause of AP and correlates with increased severity (Yang and McNabb-Baltar, 2020), raising the question whether there could be HTG associated AP in the study setting. Protein concentration decreased with increased severity in this study confirming that AP promotes protein catabolism, which involves the breakdown of proteins into amino acids (Singh et al., 2012). Hence, the presence of pancreatic inflammation could lead to an alteration in the level of metabolites (Xiao et al., 2017).

This study confirms that inflammation plays a vital role in AP severity (Iyer et al., 2020). AP patients with CRP > 150 mg/L and Albumin < 35 g/L were more likely to die in-hospital or have organ dysfunction. Furthermore, GlycA was most elevated in MSAP with a slight increase in the SAP patients, suggesting increased inflammation levels in MSAP compared to MAP and SAP. Although the differences in GlycA between the groups in this study were not significantly different, it is notable that in a recent study, Shah and colleagues show that GlycA could be used as a novel diagnostic marker of inflammation in AP when compared to healthy controls (Shah et al., 2023). Increases in GlycA and GlycB have been shown in other acute phase inflammatory diseases such as COVID-19 (Holmes et al., 2021; Lodge et al., 2021). Here, we show that GlycA could be targeted as a biomarkers of inflammation in AP.

The transcription factor, NF-_k_B, has been shown to promote the expression of genes involved in inflammation, which are vital in various AP stages (Rakonczay et al., 2008). AP is an immune disorder characterized by the release of either anti or proinflammatory cytokines (Zhang et al., 2017). The impaired anti-inflammatory function might be linked to reduced HDL-C levels in AP leading to increased free fatty acids (FFAs), which produce an acidic microenvironment (Zhang et al., 2017). HDL-C also possesses an antioxidant ability, which is mediated by the paraoxonase-1 activity in AP (Yuksekdag et al., 2019). Notably, the presence of FFAs results from the breakdown of TG including chylomicrons and is associated with HTG AP. The TGs are inherently not toxic but the resultant lipotoxicity from the FFAs causes the release of intracellular calcium resulting in acinar cell injury (Yang and McNabb-Baltar, 2020).

### Study limitations

“Sick” control samples of patients presenting with other inflammatory diseases that could result in the production of similar metabolites or lipoprotein profiles were not included in this study, hence the specificity of the metabolic changes could not be fully ascertained. However, given that metabolites in different diseases could be different due to differences in the disease course, metabolomics testing stands to differentiate these markers due to the differing specificities and the associated time bound nature (Gu and Tong, 2019). This study is a pilot study with small samples sizes (30 AP patients), but comparatively, similar to other high profile published work and findings.

#### Conclusion

This study provides crucial information for subsequent validation studies on a larger cohort. These preliminary findings point to the possibility of using metabolites and lipoproteins in delineating AP severity which adds value in understanding the pathophysiological conditions of this disease, hence ultimately leading to early diagnosis and better care.

## Conflict of interest

The authors declare that the research was conducted in the absence of any commercial or financial relationships that could be construed as a potential conflict of interest.

## Author contributions

JM: Sample acquisition/processing, analysis, interpretation, writing-original draft, writing-review and editing. NE: formal analysis, interpretation, writing-original draft, writing-review and editing. AW: design/methodology, formal analysis, interpretation, writing-review and editing. OM: clinical data acquisition, formal analysis, interpretation, writing-review and editing. JD: clinical data acquisition, interpretation, writing-review and editing, MD: clinical data analysis, interpretation, writing-review and editing. SC: contributed analytical tools, methodology data analysis, interpretation, writing-review and editing. PF: conceptualisation, obtained funding, formal analysis and interpretation, project administration, supervision, writing-review and editing. All authors approved the final version.

## Funding

This study was supported by the South African National Research Foundation Grant 121277 and the University of the Witwatersrand Faculty Research Committee Individual Grant 001254844110151211055254.

## Data Availability

All data produced in the present study are available upon reasonable request to the authors

## Acknowledgements

The patients, clinical staff, and clerks at Chris Hani Baragwanath Academic Hospital, Hepatopancreatobiliary Unit and the Department of Surgery of the University of the Witwatersrand are acknowledged. The Human Metabolomics Centre at the North-West University for NMR Spectroscopy studies.

## Supplementary material

**Tables S1-S10** where **Table S1** is the table of quantified signals and their relative assignment and multiplicity; **Table S2** is the Aetiology and clinical characteristics of the AP patients, **Table S3**: Clinical tests of patients with acute pancreatitis of different severity groups; **Table S4**. Comparison of biochemical tests for the different aetiologies of acute pancreatitis; **Table S5**. Clinical tests of patients with acute pancreatitis with biliary aetiology. All values are represented as median; **Table S6:** Clinical tests of patients with alcohol-induced AP, values are represented as median; **Table S7:** Comparison of metabolites with different aetiology groups of Acute Pancreatitis; **Table S8:** Correlation of metabolites with inverted first dimension of KODAMA. Significantly dysregulated metabolites and lipids; **Table S9:** Spearman correlation between lipoprotein parameters and inverted first dimension of KODAMA and **Table S10:** Comparison of Metabolites and lipids with severity in Acute Pancreatitis.

## Data availability statement

All data produced in the present study are available upon reasonable request to the authors.

## Notes

### Competing Interest Statement

The authors have declared no competing interest.

### Author Declarations

Ethics clearance for this study was obtained from the Human Research Ethics Committee (Medical) of the University of the Witwatersrand (Ethics No. M190407).

