## Supplementary Tables for "Metabolites and lipoproteins may predict the severity of early Acute Pancreatitis in a South African cohort"

**Table S1.** List of the quantified signals and their relative assignment and multiplicity.

| **Metabolite** | **Assignment (multiplicity)** |
| --- | --- |
| Formate | 8.45 (s) |
| Unknown signal at 8.12 ppm | 8.12 (d) |
| Unknown signal at 8.07 ppm | 8.07 (d) |
| Phenylalanine | 7.42 (m) |
| Tyrosine | 7.19 (m) |
| Unknown signal at 7.14 ppm | 7.14 (m) |
| Histidine | 7.05 (d) |
| Glucose | 5.23 (d) |
| Mannose | 5.18 (d) |
| Unknown signal at 5.15 ppm | 5.15 (d) |
| Unknown signal at 5.09 ppm | 5.09 (d) |
| Unknown signal at 5.01 ppm | 5.01 (d) |
| Ascorbate | 4.50 (d) |
| Threonine | 4.24 (m) |
| Lactate | 4.11 (q) |
| Creatinine | 4.05 (s) |
| Creatine | 3.92 (s) |
| Glycine | 3.55 (s) |
| Methanol | 3.35 (s) |
| Unknown signal at 2.55 ppm | 2.55 (s) |
| Citrate | 2.53 (d) |
| Glutamine | 2.45 (m) |
| Pyruvate | 2.36 (s) |
| Glutamate | 2.53 (m) |
| Acetoacetate | 2.22 (s) |
| Acetate | 1.91 (s) |
| Alanine | 1.47 (d) |
| Unknown signal at 1.45 ppm | 1.45 (d) |
| Unknown signal at 1.43 ppm | 1.43 (d) |
| 3-Hydroxybutyrate | 1.19 (d) |
| Ethanol | 1.17 (t) |
| Unknown signal at 1.16 ppm | 1.16 (d) |
| Unknown signal at 1.14 ppm | 1.14 (d) |
| Unknown signal at 1.11 ppm | 1.11 (d) |
| Unknown signal at 1.06 ppm | 1.06 (d) |
| Valine | 1.04 (d) |
| Isoleucine | 1.00 (d) |
| Leucine | 0.95 (dd) |
| 2-Hydroxybutyrate | 0.89 (t) |
| Protein NH | 10.00-6.00 |
| Unsaturated lipid -CH=CH- | 5.50-5.10 |
| Lipid alpha-CH2 | 2.25-2.15 |
| Cholesterol | 0.70-0.60 |
| Lipid =CH-CH2-CH= | 2.85-2.65 |
| Glycerol phospholipid | 4.08-4.03 |
| Phospholipid | 3.68-3.62 |
| Lipid beta-CH2 | 1.65-1.40 |
| Lipid CH2 | 1.40-1.10 |
| Lipid CH3 | 1.10-1.08 |
| GlycB | 2.07 (m) |
| GlycA | 2.03 (m) |

s- singlet, d - doublet, dd - doublet of doublets, m - multiplet, q- quartet, t- triplet

**Table S2**. Aetiology and clinical characteristics of the AP patients

| **Feature** | **Biliary (N=12)** | **Alcohol (N=16)** | **ARVs (N=2)** | **p-value** |
| --- | --- | --- | --- | --- |
| BMI, median [IQR] | 39.8 [34.4 43.4] | 26.4 [23.7 29.5] | 22.6 [22.6 22.6] | 0.0134 |
| Age, median [IQR] | 47.5 [37.2 58] | 36.5 [30.8 44.5] | 50 [46 54] | 0.0893 |
| Gender |  |  |  | 0.0129 |
| F, n (%) | 9 (75.0) | 4 (25.0) | 2 (100.0) |  |
| M, n (%) | 3 (25.0) | 12 (75.0) | 0 (0.0) |  |
| Days of hospitalization, median [IQR] | 10.5 [8.8 13.5] | 8 [4.8 9.5] | 7 [4 10] | 0.203 |
| Renovascular disease | |  |  | 0.402 |
| N, n (%) | 10 (83.3) | 14 (87.5) | 1 (50.0) |  |
| Y, n (%) | 2 (16.7) | 2 (12.5) | 1 (50.0) |  |
| Diabetic |  |  |  | <0.001 |
| N, n (%) | 12 (100.0) | 15 (93.8) | 2 (100.0) |  |
| Y, n (%) | 0 (0.0) | 1 (6.2) | 0 (0.0) |  |
| HIV |  |  |  | 0.188 |
| N, n (%) | 11 (91.7) | 12 (75.0) | 1 (50.0) |  |
| Y, n (%) | 1 (8.3) | 4 (25.0) | 1 (50.0) |  |
| Organ dysfunction | |  |  | 0.250 |
| no, n (%) | 8 (66.7) | 10 (62.5) | 1 (50.0) |  |
| renal, n (%) | 0 (0.0) | 1 (6.2) | 0 (0.0) |  |
| respiratory, n (%) | 2 (16.7) | 5 (31.2) | 0 (0.0) |  |
| transient renal, n (%) | 2 (16.7) | 0 (0.0) | 1 (50.0) |  |
| Local Complications | |  |  | <0.001 |
| N, n (%) | 10 (83.3) | 11 (78.6) | 2 (100.0) |  |
| Y, n (%) | 2 (16.7) | 3 (21.4) | 0 (0.0) |  |
| Admission to ICU | |  |  | 0.590 |
| N, n (%) | 10 (83.3) | 13 (81.2) | 1 (50.0) |  |
| Y, n (%) | 2 (16.7) | 3 (18.8) | 1 (50.0) |  |
| Surgical.procedures | |  |  | 0.0481 |
| cholecystectomy, n (%) | 3 (27.3) | 0 (0.0) | 0 (0.0) |  |
| ERCP, n (%) | 2 (18.2) | 0 (0.0) | 0 (0.0) |  |
| None, n (%) | 6 (54.5) | 14 (100.0) | 2 (100.0) |  |
| hospital death | |  |  | 0.291 |
| no, n (%) | 11 (91.7) | 13 (81.2) | 1 (50.0) |  |
| yes, n (%) | 1 (8.3) | 3 (18.8) | 1 (50.0) |  |

ARVs – Antiretrovirals, BMI – Body mass index, HIV - human immunodeficiency virus, ICU – Intensive care unit, ERCP - Endoscopic retrograde cholangiopancreatography, IQR- Interquartile range.

**Table S3.** Clinical tests of patients with acute pancreatitis of different severity groups

| **Features** | **Physiological range** | **MAP, median**  **(N=8)** | **MSAP, median**  **(N=14)** | **SAP, median**  **(N=8)** | ***p*-value** | **FDR** |
| --- | --- | --- | --- | --- | --- | --- |
| Amylase (U/L) | 28- 110 | 1009 | 652 | 794 | 0.820 | 0.865 |
| Lipase (U/L) | 13-60 | 649 | 760 | 1170 | 0.640 | 0.846 |
| WBC (10x^9/L) | 3.92-10.40 | 10.8 | 12.75 | 8.2 | 0.538 | 0.846 |
| Haemoglobin (g/dL) | 13.4- 17.5 | 12.8 | 14.35 | 13.95 | 0.328 | 0.846 |
| Haematocrit, (L/L) | 0.390 - 0.510 | 0.35 | 0.4 | 0.4 | 0.142 | 0.846 |
| Platelets (10x^9/L) | 171-388 | 230 | 227.5 | 224.5 | 0.668 | 0.846 |
| Na (mmol/L) | 136- 145 | 136 | 136 | 137 | 0.618 | 0.846 |
| K (mmol/L) | 3.5- 5.1 | 4.2 | 4.3 | 4.5 | 0.523 | 0.846 |
| Cl (mmol/L) | 98-107 | 96 | 100 | 85 | 0.578 | 0.846 |
| Urea (mmol/L) | 2.1-7.1 | 4 | 8.75 | 9.6 | 0.364 | 0.846 |
| CRP Reactive protein (mg/L) | <10 | 83 | 115 | 232 | 0.793 | 0.865 |
| **Liver function test** |  |  |  |  |  |  |
| Total protein, (g/L) | 60-78 | 71.5 | 69 | 62 | 0.391 | 0.846 |
| Albumin, (g/L) | 35-52 | 37 | 35.5 | 34.5 | 0.391 | 0.846 |
| Total bilirubin, (µmol/L) | 5-21 | 20 | 17.5 | 48.5 | 0.400 | 0.846 |
| Conjugated bilirubin (µmol/L) | 0-3 | 12 | 5.5 | 31.5 | 0.468 | 0.846 |
| ALT (U/L) | 10-40 | 34 | 33.5 | 102.5 | 0.509 | 0.846 |
| AST (U/L) | 15-40 | 35 | 36 | 164 | 0.102 | 0.846 |
| ALP (U/L) | 53-128 | 91 | 111 | 144 | 0.901 | 0.901 |
| GGT(U/L) | < 68 | 375 | 119 | 397 | 0.749 | 0.865 |
| FDR – False discovery rate; WBC – White blood count, CRP: C-reactive protein; Alanine transaminase (ALT), Aspartate transaminase (AST), Alkaline phosphatase (ALP), Gamma glutamyl transferase (GGT), FDR – False discovery rate, IQR- Interquartile range, (U/L) - Units per litre, L – litre, (g/dL) - grams per decilitre, (mmol/L) - Millimoles per litre, (mg/L) - milligrams per litre, , (g/L) - grams per litre, , (µmol/L) - micromole per litre | | | | | | |

**Table S4**. Comparison of biochemical tests for the different aetiologies of acute pancreatitis

| **Feature** | **Biliary, median [IQR] (N=12)** | **Alcohol, median [IQR] (N=16)** | **ARVs, median [IQR] (N=2)** | **p-value** | **FDR** |
| --- | --- | --- | --- | --- | --- |
| Amylase (U/L) | 1914 [1696.5 2642] | 450 [370.5 794] | 450 [450 450] | 0.001 | 0.008 |
| Lipase (U/L) | 1129.5 [529.5 1678.25] | 749 [489.75 1031] | NA | 0.339 | 0.496 |
| WBC (10x^9/L) | 13.1 [10.9 14.7] | 8.05 [6.875 10.875] | 12.1 [8.95 15.25] | 0.032 | 0.154 |
| Haemoglobin (g/dL) | 13.4 [11.2 14.8] | 13.95 [12.675 15.25] | 12.55 [11.525 13.575] | 0.733 | 0.871 |
| Haematocrit, (L/L) | 0.4 [0.325 0.475] | 0.4 [0.35 0.4] | 0.4 [0.35 0.45] | 0.914 | 0.961 |
| Platelets (10x^9/L) | 235 [212 303.5] | 209.5 [133.75 252.75] | 264 [256 272] | 0.183 | 0.387 |
| Na (mmol/L) | 140 [135 140] | 134.5 [129 137] | 137 [137 137] | 0.240 | 0.432 |
| K (mmol/L) | 4.4 [4 27.375] | 4.4 [3.95 5.175] | 3.2 [3.2 3.2] | 0.325 | 0.496 |
| Cl (mmol/L) | 54 [29.2 83.5] | 100 [96 101] | 97 [97 97] | 0.570 | 0.774 |
| Urea (mmol/L) | 7.5 [5.8 8.8] | 8.5 [2.9 12.45] | 8.7 [8.7 8.7] | 0.961 | 0.961 |
| CRP (mg/L) | 46 [32 136] | 162.5 [70.25 297.25] | 337 [337 337] | 0.145 | 0.344 |
| Total. Protein (g/L) | 66.5 [60.75 71.25] | 66 [57.5 72] | 62 [62 62] | 0.908 | 0.961 |
| Albumin (g/L) | 36 [35 38.5] | 35 [31 41] | 29.5 [24.75 34.25] | 0.715 | 0.871 |
| Total Bilirubin (µmol/L) | 42.5 [15.5 91.75] | 15 [8 21] | 31 [22 40] | 0.250 | 0.432 |
| DBil. (µmol/L) | 21.5 [11.25 57.5] | 5 [4 6] | 26 [16.5 35.5] | 0.077 | 0.209 |
| ALT (U/L) | 160 [82.5 232] | 23 [22 37] | 27 [22 32] | 0.013 | 0.085 |
| AST (U/L) | 91.5 [44.25 231.75] | 33 [27.25 46.25] | 50 [42 58] | 0.074 | 0.209 |
| ALP (U/L) | 267 [231.25 304] | 75 [64.25 90.25] | 93.5 [84.75 102.25] | <0.001 | 0.003 |
| GGT (U/L) | 468 [328.5 1028.25] | 175.5 [65.25 317.75] | 192 [117.5 266.5] | 0.053 | 0.200 |
| FDR: false discovery rate; WBC – White blood count, CRP: C-reactive protein; Alanine transaminase (ALT), Aspartate transaminase (AST), Alkaline phosphatase (ALP), Gamma glutamyl transferase (GGT), FDR – False discovery rate, IQR- Interquartile range, (U/L) - Units per litre, L – litre, (g/dL) - grams per decilitre, (mmol/L) - Millimoles per litre, (mg/L) - milligrams per litre, , (g/L) - grams per litre, , (µmol/L) - micromole per litre | | | | | |

**Table S5**. Clinical tests of patients with acute pancreatitis with biliary aetiology. All values are represented as median [interquartile range]

| **Feature** | **Mild (N=4)** | **Moderate (N=5)** | **Severe (N=3)** | **rho** | **p-value** | **FDR** |
| --- | --- | --- | --- | --- | --- | --- |
| Amylase (U/L) | 1710.5 [1486.5 2193.75] | 2604 [1993.5 3840.75] | 1914 [1205 2038.5] | 0.05 | 0.877 | 0.987 |
| Lipase (U/L) | 1294 [797 1791] | 847.5 [471.25 1566.75] | 1322.5 [1246.25 1398.75] | 0.15 | 0.715 | 0.933 |
| WBC (10x^9/L) | 11 [10.9 13.5] | 14 [13.1 15.2] | 8.4 [8.2 11.3] | -0.23 | 0.489 | 0.909 |
| Haemoglobin (g/dL) | 10 [10 10] | 13.4 [11.2 14.2] | 16.1 [14.75 16.25] | 0.71 | 0.031 | 0.325 |
| Haematocrit, (L/L) | 0.34 [0.337 0.344] | 0.408 [0.355 0.443] | 0.45 [0.414 0.496] | 0.66 | 0.036 | 0.325 |
| Platelets (10x^9/L) | 323 [276.5 332.5] | 242 [210 284] | 214 [158 224.5] | -0.51 | 0.112 | 0.505 |
| Na (mmol/L) | 140 [71.75 142] | 139 [135 139.5] | 140 [140 140] | -0.04 | 0.932 | 0980 |
| K (mmol/L) | 35 [18.95 71] | 4.1 [4 4.2] | 4.5 [4.5 4.5] | -0.19 | 0.725 | 0.933 |
| Cl (mmol/L) | NA | NA | NA | NA | NA | NA |
| Urea (mmol/L) | NA | NA | NA | NA | NA | NA |
| Creatinine (mg/dL) | 53.5 [50.75 56.25] | 107 [87.5 113.75] | 182 [139.5 224.5] | 0.54 | 0.167 | 0.516 |
| CRP (mg/L) | 60 [48.5 71.5] | 46 [32 136] | 123.5 [69.25 177.75] | 0 |  |  |
| Total protein (g/L) | 72 [72 72] | 65.5 [59.5 78.5] | 62 [61.5 66.5] | -0.26 | 0.541 | 0.909 |
| Albumin (g/L) | 38 [37.5 38.5] | 35 [35 36] | 36 [34.5 38] | -0.21 | 0.556 | 0.909 |
| Total bilirubin (µmol/L) | 35 [27.5 42.5] | 14 [9 35] | 115 [95.5 117.5] | 0.55 | 0.102 | 0.505 |
| DBil, (µmol/L) | 21.5 [18.25 24.75] | 11 [2 12] | 58 [57 77] | 0.45 | 0.193 | 0.516 |
| ALT (U/L) | 208 [200.5 215.5] | 75 [38 127] | 235 [170 316.5] | 0.14 | 0.690 | 0.933 |
| AST (U/L) | 88 [78.5 97.5] | 36 [30 76] | 239 [224.5 248.5] | 0.44 | 0.201 | 0.516 |
| ALP (U/L) | 335 [300 370] | 225 [187 269] | 271 [260.5 293] | -0.05 | 0.885 | 0.987 |
| GGT (U/L) | 956 [665.5 1246.5] | 313 [119 426] | 1187 [848.5 1472] | 0.3 | 0.395 | 0.889 |
| Rho - Spearman’s correlation coefficient, FDR – False discovery rate, IQR- Interquartile range; WBC: white blood count; CRP: C-Reactive protein; DBil: Conjugated bilirubin ; ALT: Alanine transaminase; AST: Aspartate transaminase; ALP: Alkaline phosphatase; GGT: Gamma-glutamyl transferase, (U/L) - Units per litre, L – litre, (g/dL) - grams per decilitre, (mg/dL) - milligrams per decilitre (mmol/L) - Millimoles per litre, (mg/L) - milligrams per litre, , (g/L) - grams per litre, , (µmol/L) - micromole per litre | | | | | | |

**Table S6**. Clinical tests of patients with alcohol-induced AP, values are represented as median [interquartile range].

| **Feature** | **Mild (N=4)** | **Moderate (N=8)** | **Severe (N=4)** | **rho** | **p-value** | **FDR** |
| --- | --- | --- | --- | --- | --- | --- |
| Amylase (U/L) | 390 [265 620] | 418 [372 798.75] | 764 [566.5 794] | 0.21 | 0.460 | 0.932 |
| Lipase (U/L) | 649 [379.25 947.5] | 760 [686.5 926.5] | 738 [402 1068.5] | -0.01 | 0.974 | 0.974 |
| WBC (10x^9/L) | 6.55 [4.625 9.85] | 9.15 [7.1 11.35] | 8.95 [7.575 10.5] | 0.15 | 0.570 | 0.932 |
| Haemoglobin (g/dL) | 13.25 [12.675 13.875] | 15.3 [14.1 15.45] | 12.7 [11.275 13.925] | -0.12 | 0.671 | 0.932 |
| Haematocrit, (L/L) | 0.408 [0.374 0.433] | 0.438 [0.412 0.459] | 0.381 [0.342 0.417] | -0.18 | 0.521 | 0.932 |
| Platelets (10x^9/L) | 200 [177.5 219.5] | 180.5 [120.25 258] | 224 [187.75 279.25] | 0.13 | 0.620 | 0.932 |
| Na (mmol/L) | 132 [99.225 136.25] | 134.5 [132.5 136.5] | 134.5 [100.15 138.25] | 0.13 | 0.659 | 0.932 |
| K (mmol/L) | 4.15 [3.975 4.25] | 5.05 [4.05 5.375] | 4.45 [4.275 22.625] | 0.35 | 0.217 | 0.932 |
| Cl (mmol/L) | 96.5 [96.25 96.75] | 100 [95.75 102.75] | 101 [85 319.5] | 0.29 | 0.455 | 0.932 |
| Urea (mmol/L) | 6.25 [5.125 7.375] | 9.2 [5.35 34.6] | 8.75 [5.275 12.225] | 0.09 | 0.840 | 0.932 |
| Creatinine (mg/dL) | 99.5 [81 112.5] | 65 [64 134] | 75.5 [69.25 252] | -0.05 | 0.865 | 0.932 |
| CRP (mg/L) | 191.5 [78.75 318.5] | 119 [46.75 247.5] | 222 [155.75 261.75] | -0.05 | 0.874 | 0.932 |
| Total protein (g/L) | 71 [62 71.5] | 69.5 [61 72] | 61 [53.5 63] | -0.28 | 0.387 | 0.932 |
| Albumin (g/L) | 35 [33 39.5] | 40.5 [34 42.5] | 31 [29.5 33.5] | -0.33 | 0.264 | 0.932 |
| Total bilirubin (µmol/L) | 11 [10 16] | 17.5 [9.75 20.75] | 14 [6.5 47.75] | -0.04 | 0.885 | 0.932 |
| DBil, (µmol/L) | 5 [4.5 8.5] | 4.5 [4 5.75] | 5 [3.5 33.5] | -0.05 | 0.868 | 0.932 |
| ALT (U/L) | 23 [22.5 28.5] | 22 [19.5 28.25] | 68.5 [33.25 118.5] | 0.37 | 0.217 | 0.932 |
| AST (U/L) | 29 [26.5 32] | 31 [27.5 42] | 79.5 [33.5 164.75] | 0.37 | 0.199 | 0.932 |
| ALP (U/L) | 88 [87.5 89.5] | 64 [56.5 66] | 104 [75 138.5] | 0.11 | 0.708 | 0.932 |
| GGT (U/L) | 319 [181.5 347.5] | 81 [57.5 203] | 271.5 [216.5 356] | 0.1 | 0.733 | 0.932 |
| Rho - Spearman’s correlation coefficient, FDR – False discovery rate, IQR- Interquartile range; WBC: white blood count; CRP: C-Reactive protein; DBil: Conjugated bilirubin ; ALT: Alanine transaminase; AST: Aspartate transaminase; ALP: Alkaline phosphatase; GGT: Gamma-glutamyl transferase, (U/L) - Units per litre, L – litre, (g/dL) - grams per decilitre, (mg/dL) - milligrams per decilitre (mmol/L) - Millimoles per litre, (mg/L) - milligrams per litre, , (g/L) - grams per litre, , (µmol/L) - micromole per litre | | | | | | |

**Table S7:** Comparison of metabolites with different aetiology groups of Acute Pancreatitis

| **Metabolites** | **Biliary, median [IQR]**  **(N=12)** | **Alcohol, median [IQR]**  **(N=16)** | **ARVs, median [IQR]**  **(N=2)** | **p-value** | **FDR** |
| --- | --- | --- | --- | --- | --- |
| Formate | 0.016 [0.012 0.02] | 0.015 [0.01 0.017] | 0.01 [0.009 0.011] | 0.462 | 0.760 |
| Unknown signal at 8.12 ppm | 0.006 [0 0.012] | 0.003 [0 0.009] | 0.006 [0.003 0.008] | 0.811 | 0.871 |
| Unknown signal at 8.07 ppm | 0 [0 0] | 0 [0 0.001] | 0 [0 0] | 0.730 | 0.871 |
| Phenylalanine | 0.147 [0.106 0.158] | 0.129 [0.121 0.15] | 0.279 [0.218 0.341] | 0.218 | 0.760 |
| **Tyrosine** | **0.037 [0.033 0.04]** | **0.054 [0.045 0.058]** | **0.049 [0.046 0.051]** | **0.005** | **0.203** |
| Unknown signal at 7.14 ppm | 0.007 [0 0.041] | 0 [0 0.024] | 0.019 [0.009 0.028] | 0.806 | 0.871 |
| Histidine | 0.088 [0.065 0.095] | 0.073 [0.063 0.083] | 0.096 [0.085 0.108] | 0.522 | 0.760 |
| Glucose | 3.138 [2.846 4.186] | 3.399 [2.324 3.909] | 2.53 [2.451 2.609] | 0.348 | 0.760 |
| Mannose | 0.058 [0.045 0.065] | 0.066 [0.047 0.098] | 0.082 [0.071 0.093] | 0.398 | 0.760 |
| Unknown signal at 5.15 ppm | 0 [0 0] | 0 [0 0] | 0 [0 0] | 0.472 | 0.760 |
| Unknown signal at 5.09 ppm | 0 [0 0.025] | 0 [0 0] | 0.011 [0.006 0.017] | 0.351 | 0.760 |
| Unknown signal at 5.01 ppm | 0 [0 0] | 0 [0 0] | 0 [0 0] | 0.472 | 0.760 |
| Ascorbate | 0 [0 0.002] | 0 [0 0.01] | 0 [0 0] | 0.516 | 760 |
| Threonine | 0.072 [0.045 0.096] | 0.061 [0.03 0.076] | 0 [0 0] | 0.126 | 0.584 |
| Lactate | 2.861 [2.367 4.054] | 3.022 [2.291 3.63] | 3.811 [3.289 4.334] | 0.707 | 0.871 |
| Creatinine | 0.072 [0.063 0.089] | 0.072 [0.054 0.1] | 0.175 [0.137 0.213] | 0.261 | 0.760 |
| Creatine | 0.032 [0.012 0.07] | 0.058 [0.02 0.11] | 0.138 [0.075 0.2] | 0.633 | 0.852 |
| Glycine | 0.312 [0.27 0.354] | 0.271 [0.227 0.31] | 0.335 [0.333 0.338] | 0.273 | 0.760 |
| Methanol | 0.067 [0.035 0.101] | 0.039 [0.025 0.051] | 0.041 [0.04 0.043] | 0.348 | 0.760 |
| Unknown signal at 2.55 ppm | 0 [0 0] | 0 [0 0] | 0.032 [0.023 0.04] | 0.009 | 0.203 |
| Citrate | 0.09 [0.046 0.121] | 0.07 [0.062 0.084] | 0 [0 0] | 0.107 | 0.561 |
| Glutamine | 0.269 [0.249 0.284] | 0.29 [0.239 0.329] | 0.306 [0.293 0.32] | 0.617 | 0.852 |
| Pyruvate | 0.064 [0.044 0.094] | 0.064 [0.038 0.106] | 0.161 [0.14 0.181] | 0.144 | 0.611 |
| Glutamate | 0.166 [0.121 0.209] | 0.169 [0.152 0.211] | 0.198 [0.197 0.199] | 0.652 | 0.852 |
| **Acetoacetate** | **0.073 [0.04 0.112]** | **0.193 [0.116 0.415]** | **0.182 [0.119 0.244]** | **0.012** | **0.203** |
| Acetate | 0.032 [0.024 0.034] | 0.028 [0.024 0.033] | 0.033 [0.032 0.033] | 0.837 | 0.871 |
| Alanine | 0.355 [0.32 0.434] | 0.341 [0.307 0.365] | 0.42 [0.399 0.441] | 0.417 | 0.760 |
| Unknown signal at 1.45 ppm | 0.053 [0.044 0.065] | 0.056 [0.041 0.079] | 0.074 [0.059 0.089] | 0.785 | 0.871 |
| Unknown signal at 1.43 ppm | 0 [0 0] | 0 [0 0] | 0 [0 0] | 0.472 | 0.760 |
| **3-Hydroxybutyrate** | **0.086 [0.051 0.203]** | **0.435 [0.13 1.692]** | **0.244 [0.164 0.324]** | **0.055** | **0.478** |
| Ethanol | 0.006 [0 0.014] | 0 [0 0] | 0.038 [0.019 0.058] | 0.110 | 0.561 |
| Unknown signal at 1.16 ppm | 0.039 [0 0.096] | 0.126 [0 5.133] | 0.767 [0.384 1.151] | 0.514 | 0.760 |
| Unknown signal at 1.14 ppm | 0.022 [0 0.039] | 0 [0 0.021] | 0.148 [0.106 0.191] | 0.075 | 0.478 |
| Unknown signal at 1.11 ppm | 0.077 [0.059 0.083] | 0.086 [0.073 0.104] | 0.07 [0.066 0.074] | 0.157 | 0.617 |
| Unknown signal at 1.06 ppm | 0.042 [0.032 0.05] | 0.053 [0.036 0.094] | 0.064 [0.051 0.076] | 0.377 | 0.760 |
| **Valine** | **0.178 [0.155 0.218]** | **0.237 [0.21 0.268]** | **0.236 [0.206 0.265]** | **0.059** | **0.478** |
| Isoleucine | 0.038 [0.027 0.054] | 0.059 [0.05 0.068] | 0.072 [0.064 0.08] | 0.420 | 0.478 |
| Leucine | 0.072 [0.062 0.099] | 0.099 [0.083 0.11] | 0.104 [0.094 0.115] | 0.066 | 0.478 |
| 2-Hydroxybutyrate | 0.011 [0 0.079] | 0.072 [0.02 0.099] | 0.062 [0.038 0.085] | 0.367 | 0.760 |
| Protein NH | 133.86 [119.887 139.57] | 130.264 [119.303 139.803] | 149.873 [138.618 161.129] | 0.701 | 0.871 |
| Unsaturated lipid -CH=CH- | 10.76 [9.163 13.55] | 9.782 [8.757 13.015] | 10.199 [10.137 10.261] | 0.955 | 0.955 |
| Lipid alpha-CH2 | 1.808 [1.095 3.333] | 2.19 [1.73 2.879] | 1.573 [1.534 1.613] | 0.517 | 0.760 |
| Cholesterol | 0.911 [0.514 1.015] | 0.639 [0.586 0.734] | 0.843 [0.768 0.918] | 0.234 | 0.760 |
| Lipid =CH-CH2-CH= | 5.073 [3.569 5.666] | 4.309 [3.946 5.495] | 4.826 [4.748 4.903] | 0.821 | 0.871 |
| Glycorol phospholipid | 0.416 [0.122 0.978] | 0.504 [0.249 0.848] | 0.18 [0.098 0.262] | 0.648 | 0.852 |
| Phospholipid | 3.675 [3.353 4.714] | 3.602 [3.346 4.345] | 4.881 [4.482 5.281] | 0.362 | 0.760 |
| Lipid beta-CH2 | 6.904 [6.076 8.183] | 5.872 [5.297 8.456] | 6.659 [6.554 6.764] | 0.737 | 0.871 |
| Lipid CH2 | 64.754 [50.366 75.038] | 62.248 [53.163 83.574] | 59.558 [58.656 60.461] | 0.881 | 0.899 |
| Lipid CH3 | 25.489 [21.931 29.467] | 23.373 [19.958 27.419] | 25.432 [25.227 25.637] | 70.78 | 0.871 |
| GlycB | 0.374 [0.364 0.48] | 0.412 [0.367 0.467] | 0.56 [0.505 0.614] | 0.286 | 0.760 |
| GlycA | 1.719 [1.503 2.005] | 1.913 [1.59 2.185] | 2.62 [2.287 2.953] | 0.316 | 0.760 |

ARVs- antiretrovirals, FDR – False discovery rate, IQR- Interquartile range

**Table S8.** Correlation of metabolites with inverted first dimension of KODAMA. Significantly dysregulated metabolites and lipids are represented in bold.

| **Feature** | **rho** | **p-value** | **FDR** |
| --- | --- | --- | --- |
| Formate | -0.05 | 0.776 | 0.842 |
| Unknown signal at 8.12 ppm | -0.11 | 0.531 | 0.639 |
| Unknown signal at 8.07 ppm | 0.05 | 0.747 | 0.828 |
| **Phenylalanine** | **-0.51** | **0.001** | **0.005** |
| Tyrosine | 0.26 | 0.116 | 0.211 |
| Unknown signal at 7.14 ppm | -0.31 | 0.062 | 0.138 |
| Histidine | 0.19 | 0.266 | 0.357 |
| Glucose | -0.04 | 0.834 | 0.868 |
| **Mannose** | **-0.46** | **0.005** | **0.017** |
| Unknown signal at 5.15 ppm | -0.28 | 0.092 | 0.174 |
| Unknown signal at 5.09 ppm | -0.17 | 0.306 | 0.401 |
| Unknown signal at 5.01 ppm | -0.28 | 0.092 | 0.174 |
| **Ascorbate** | **0.44** | **0.006** | **0.020** |
| **Threonine** | **0.35** | **0.032** | **0.0898** |
| **Lactate** | **-0.62** | **<0.001** | **<0.001** |
| Creatinine | 0.01 | 0.934 | 0.934 |
| Creatine | -0.31 | 0.062 | 0.138 |
| Glycine | 0.02 | 0.928 | 0.934 |
| **Methanol** | **0.39** | **0.018** | **0.053** |
| Unknown signal at 2.55 ppm | -0.2 | 0.229 | 0.341 |
| Citrate | 0.2 | 0.242 | 0.341 |
| **Glutamine** | **0.57** | **<0.001** | **0.002** |
| **Pyruvate** | **-0.33** | **0.046** | **0.110** |
| Glutamate | -0.16 | 0.329 | 0.420 |
| **Acetoacetate** | **-0.53** | **<0.001** | **0.004** |
| Acetate | -0.13 | 0.425 | 0.529 |
| Alanine | 0.21 | 0.216 | 0.333 |
| Unknown signal at 1.45 ppm | -0.45 | 0.005 | 0.018 |
| Unknown signal at 1.43 ppm | -0.28 | 0.092 | 0.174 |
| **3-Hydroxybutyrate** | **-0.35** | **0.034** | **0.090** |
| **Ethanol** | **0.63** | **<0.001** | **<0.001** |
| Unknown signal at 1.16 ppm | -0.1 | 0.566 | 0.656 |
| Unknown signal at 1.14 ppm | -0.2 | 0.242 | 0.341 |
| Unknown signal at 1.11 ppm | -0.52 | <0.000 | 0.004 |
| Unknown signal at 1.06 ppm | -0.33 | 0.044 | 0.110 |
| Valine | 0.22 | 0.200 | 0.320 |
| Isoleucine | 0.04 | 0.834 | 0.868 |
| Leucine | -0.1 | 0.538 | 0.639 |
| **2-Hydroxybutyrate** | **-0.52** | **<0.001** | **0.004** |
| **Protein NH** | **0.82** | **<0.001** | **<0.001** |
| Unsaturated lipid -CH=CH- | 0.65 | 0.000 | 0.000 |
| Lipid alpha-CH2 | -0.28 | 0.089 | 0.174 |
| **Cholesterol** | **0.62** | **<0.001** | **<0.001** |
| **Lipid =CH-CH2-CH=** | **0.74** | **<0.001** | **<0.001** |
| Glycorol phospholipid | 0.07 | 0.695 | 0.788 |
| Phospholipid | 0.22 | 0.199 | 0.320 |
| **Lipid beta-CH2** | **0.77** | **<0.001** | **<0.001** |
| Lipid CH2 | 0.19 | 0.247 | 0.341 |
| **Lipid CH3** | **0.81** | **<0.001** | **<0.001** |
| GlycB | -0.23 | 0.176 | 0.309 |
| GlycA | -0.22 | 0.189 | 0.320 |

Rho - Spearman’s correlation coefficient, FDR – False discovery rate

**Table S9.** Spearman correlation between lipoprotein parameters and inverted first dimension of KODAMA. Significantly dysregulated lipids are represented in bold.

| **Feature** | **rho** | **p-value** | **FDR** |
| --- | --- | --- | --- |
| VLDL-C | 0.26 | 0.116 | 0.216 |
| **IDL-C** | **0.32** | **0.050** | **0.131** |
| **LDL-C** | **-0.39** | **0.018** | **0.059** |
| **HDL-C** | **-0.63** | **<0.001** | **<0.001** |
| VLDL-TG | 0.23 | 0.171 | 0.278 |
| IDL-TG | 0.26 | 0.114 | 0.216 |
| LDL-TG | 0.21 | 0.204 | 0.294 |
| HDL-TG | 0.22 | 0.185 | 0.283 |
| VLDL-P (nmol/L) | 0.25 | 0.135 | 0.234 |
| Large VLDL-P (nmol/L) | 0.26 | 0.113 | 0.216 |
| Medium VLDL-P (nmol/L) | 0.14 | 0.409 | 0.466 |
| Small VLDL-P (nmol/L) | 0.27 | 0.111 | 0.216 |
| **LDL-P (nmol/L)** | **-0.4** | **0.015** | **0.059** |
| Large LDL-P (nmol/L) | -0.18 | 0.283 | 0.369 |
| Medium LDL-P (nmol/L) | -0.18 | 0.274 | 0.369 |
| **Small LDL-P (nmol/L)** | **-0.47** | **0.004** | **0.019** |
| **HDL-P (µmol/L)** | **-0.65** | **<0.001** | **<0.001** |
| Large HDL-P (µmol/L) | 0.02 | 0.895 | 0.895 |
| Medium HDL-P (µmol/L) | -0.08 | 0.650 | 0.676 |
| **Small HDL-P (µmol/L)** | **-0.68** | **<0.001** | **<0.001** |
| VLDL-Z (nm) | 0.1 | 0.542 | 0.587 |
| **LDL-Z (nm)** | **0.34** | **0.0392** | **0.113** |
| **HDL-Z (nm)** | **0.68** | **<0.001** | **<0.001** |
| **Non-HDL-P (nmol/L)** | **-0.4** | **0.016** | **0.059** |
| Total-P/HDL-P | 0.16 | 0.338 | 0.418 |
| LDL-P/HDL-P | 0.14 | 0.412 | 0.466 |

Rho - Spearman’s correlation coefficient, FDR – False discovery rate, VLDL-C – Very low-density lipoprotein cholesterol, IDL – Intermediate density lipoproteins, HDL- High density lipoprotein, HDL-P- High density lipoprotein -Particle, LDL- Low-density lipoprotein, VLDL-Very-Low Density Lipoprotein, TG - Triglyceride

**Table S10.** Comparison of Metabolites and lipids with severity in Acute Pancreatitis. Significantly dysregulated lipids are represented in bold.

| **Feature** | **Mild** | **Moderate** | **Severe** | **p-value** | **FDR** |
| --- | --- | --- | --- | --- | --- |
| Formate, median [IQR] | 0.001 [-0.001 0.002] | 0.004 [0.002 0.007] | 0.002 [-0.002 0.004] | 0.288 | 0.538 |
| Unknown signal at 8.12 ppm, median [IQR] | 0.001 [-0.003 0.005] | 0.001 [0 0.003] | 0.004 [-0.001 0.008] | 0.673 | 0.838 |
| Unknown signal at 8.07 ppm, median [IQR] | 0 [0 0] | 0 [0 0.003] | 0.002 [0 0.003] | 0.661 | 0.838 |
| Phenylalanine, median [IQR] | -0.01 [-0.015 0.004] | 0.004 [-0.01 0.015] | 0.014 [0.008 0.018] | 0.033 | 0.212 |
| Tyrosine, median [IQR] | 0.002 [-0.002 0.006] | 0 [-0.007 0.003] | -0.001 [-0.003 0.004] | 0.327 | 0.576 |
| Unknown signal at 7.14 ppm, median [IQR] | 0.021 [0 0.024] | 0 [-0.001 0.015] | -0.003 [-0.009 0] | 0.071 | 0.248 |
| Histidine, median [IQR] | -0.005 [-0.009 0.003] | -0.001 [-0.008 0.011] | 0.001 [-0.004 0.005] | 0.491 | 0.716 |
| Glucose, median [IQR] | -0.015 [-0.113 0.461] | 0.133 [-0.042 0.476] | -0.425 [-0.693 0.155] | 0.466 | 0.710 |
| Mannose, median [IQR] | -0.002 [-0.013 0.007] | 0.002 [-0.01 0.004] | 0.001 [-0.007 0.012] | 0.801 | 0.923 |
| Unknown signal at 5.15 ppm, median [IQR] | 0 [0 0] | 0 [0 0] | 0 [0 0] | 0.276 | 0.538 |
| Unknown signal at 5.09 ppm, median [IQR] | 0.001 [0 0.013] | 0 [-0.001 0.004] | 0 [0 0.004] | 0.548 | 0.740 |
| Unknown signal at 5.01 ppm, median [IQR] | 0 [0 0] | 0 [0 0] | 0 [0 0] | 0.739 | 0.898 |
| Ascorbate, median [IQR] | 0 [0 0.004] | 0 [0 0.003] | -0.004 [-0.005 0] | 0.115 | 0.311 |
| Threonine, median [IQR] | 0.007 [0.003 0.028] | -0.003 [-0.008 0.009] | 0.004 [-0.012 0.029] | 0.473 | 0.710 |
| Lactate, median [IQR] | -0.001 [-0.194 0.312] | 0.213 [-0.064 0.42] | 0.009 [-0.766 0.068] | 0.220 | 0.487 |
| Creatinine, median [IQR] | -0.001 [-0.005 0.006] | 0.001 [-0.002 0.006] | -0.011 [-0.083 0.001] | 0.204 | 0.473 |
| Creatine, median [IQR] | 0 [-0.004 0.001] | -0.011 [-0.026 0] | -0.004 [-0.036 -0.003] | 0.241 | 0.512 |
| Glycine, median [IQR] | 0.01 [-0.001 0.02] | 0.034 [0.001 0.042] | 0.026 [0.002 0.041] | 0.424 | 0.697 |
| Methanol, median [IQR] | 0.003 [-0.011 0.012] | 0.002 [-0.004 0.036] | 0.001 [-0.002 0.001] | 0.393 | 0.668 |
| Unknown signal at 2.55 ppm, median [IQR] | 0 [0 0] | 0 [0 0.006] | 0 [0 0.036] | 0.263 | 0.537 |
| Citrate, median [IQR] | -0.003 [-0.011 0.006] | -0.001 [-0.024 0.006] | -0.002 [-0.014 0.005] | 0.890 | 0.923 |
| Glutamine, median [IQR] | -0.013 [-0.039 0] | 0.026 [0.003 0.05] | 0.028 [0.012 0.032] | 0.098 | 0.311 |
| Pyruvate, median [IQR] | -0.008 [-0.022 0] | 0.004 [-0.014 0.013] | 0.005 [-0.013 0.016] | 0.470 | 0.710 |
| Glutamate, median [IQR] | 0.02 [0.003 0.046] | 0.034 [0.018 0.063] | 0.03 [-0.01 0.052] | 0.829 | 0.923 |
| Acetoacetate, median [IQR] | 0.003 [-0.007 0.007] | -0.022 [-0.067 0.031] | 0.145 [0.061 0.421] | 0.007 | 0.092 |
| Acetate, median [IQR] | -0.001 [-0.003 0.007] | 0.002 [-0.001 0.006] | 0.004 [-0.001 0.007] | 0.819 | 0.923 |
| Alanine, median [IQR] | 0.006 [-0.036 0.046] | 0.02 [-0.009 0.044] | 0.012 [-0.023 0.027] | 0.876 | 0.923 |
| Unknown signal at 1.45 ppm, median [IQR] | 0 [-0.005 0] | 0.005 [-0.001 0.012] | 0.004 [-0.007 0.017] | 0.295 | 0.538 |
| Unknown signal at 1.43 ppm, median [IQR] | 0 [0 0] | 0 [0 0] | 0 [0 0] | 1.000 | 1.000 |
| 3-Hydroxybutyrate, median [IQR] | 0.002 [-0.015 0.003] | -0.023 [-0.213 0.016] | 0.514 [0.187 0.781] | 0.012 | 0.092 |
| Ethanol, median [IQR] | 0.003 [-0.003 0.008] | 0 [0 0] | 0 [0 0.009] | 0.623 | 0.814 |
| Unknown signal at 1.16 ppm, median [IQR] | -0.003 [-0.683 0.009] | -0.012 [-0.058 0.012] | 0 [-0.448 0] | 0.866 | 0.923 |
| Unknown signal at 1.14 ppm, median [IQR] | 0 [-0.003 0.012] | -0.007 [-0.019 0.012] | 0 [0 0.015] | 0.520 | 0.737 |
| Unknown signal at 1.11 ppm, median [IQR] | -0.01 [-0.013 -0.005] | -0.002 [-0.01 0.003] | -0.002 [-0.004 0.004] | 0.162 | 0.393 |
| Unknown signal at 1.06 ppm, median [IQR] | -0.004 [-0.005 -0.001] | -0.007 [-0.012 0] | 0.007 [0.004 0.015] | 0.004 | 0.092 |
| Valine, median [IQR] | -0.009 [-0.027 0.001] | 0.002 [-0.009 0.024] | 0.02 [0.011 0.054] | 0.071 | 0.248 |
| Isoleucine, median [IQR] | 0.003 [-0.005 0.009] | 0 [-0.002 0.012] | 0.01 [0.002 0.013] | 0.551 | 0.740 |
| Leucine, median [IQR] | -0.006 [-0.011 0.001] | 0.001 [-0.005 0.011] | 0.01 [0.006 0.018] | 0.073 | 0.248 |
| 2-Hydroxybutyrate, median [IQR] | -0.009 [-0.013 -0.003] | -0.005 [-0.025 0] | 0.004 [-0.005 0.036] | 0.116 | 0.311 |
| Protein NH, median [IQR] | 1.652 [-5.597 3.752] | 1.062 [-6.484 14.877] | -2.663 [-5.193 3.942] | 0.905 | 0.923 |
| Unsaturated lipid -CH=CH-, median [IQR] | -0.226 [-0.746 -0.077] | 0.12 [-0.457 1.535] | 0.905 [0.578 1.35] | 0.108 | 0.311 |
| Lipid alpha-CH2, median [IQR] | -0.275 [-0.414 0.046] | 0.018 [-0.128 0.213] | 0.344 [-0.118 0.564] | 0.148 | 0.377 |
| **Cholesterol, median [IQR]** | **-0.026 [-0.06 0.003]** | **0.02 [-0.052 0.106]** | **0.054 [0.044 0.176]** | **0.013** | **0.092** |
| **Lipid =CH-CH2-CH=, median [IQR]** | **-0.121 [-0.286 -0.01]** | **0.093 [-0.105 0.623]** | **0.469 [0.205 0.787]** | **0.013** | **0.092** |
| Glycorol phospholipid, median [IQR] | -0.082 [-0.137 0.003] | -0.002 [-0.214 0.036] | 0 [-0.201 0.102] | 0.840 | 0.923 |
| Phospholipid, median [IQR] | -0.202 [-0.358 0.008] | 0.168 [-0.006 0.267] | 0.06 [-0.027 0.146] | 0.064 | 0.248 |
| Lipid beta-CH2, median [IQR] | -0.138 [-0.426 -0.06] | -0.056 [-0.368 0.668] | 0.453 [0.206 1.324] | 0.061 | 0.248 |
| Lipid CH2, median [IQR] | -2.675 [-5.831 -1.865] | 1.226 [-2.266 7.464] | 3.196 [1.422 7.154] | 0.068 | 0.248 |
| **Lipid CH3, median [IQR]** | **-0.014 [-1.985 0.337]** | **0.589 [-0.394 2.7]** | **1.528 [0.599 1.627]** | **0.047** | **0.248** |
| **GlycB, median [IQR]** | **0 [-0.004 0.008]** | **0.045 [0.037 0.089]** | **0.014 [-0.001 0.038]** | **0.010** | **0.092** |
| **GlycA, median [IQR]** | **-0.021 [-0.069 0.039]** | **0.24 [0.149 0.337]** | **0.01 [-0.057 0.12]** | **0.004** | **0.092** |

FDR – False discovery rate, IQR- Interquartile range

**Table S11.** Comparison of metabolites with worse clinical outcomes in Acute Pancreatitis. The ketoacidosis metabolite, 3-hydroxybutyrate which is significantly dysregulated as well as acetoacetate and creatinine are represented in bold.

| **Feature** | **FALSE** | **TRUE** | **p-value** | **FDR** |
| --- | --- | --- | --- | --- |
| Formate, median [IQR] | 0.003 [0 0.004] | -0.001 [-0.003 0.004] | 0.659 | 0.997 |
| Unknown signal at 8.12 ppm, median [IQR] | 0.001 [-0.001 0.004] | 0.007 [-0.001 0.009] | 0.395 | 0.821 |
| Unknown signal at 8.07 ppm, median [IQR] | 0 [0 0.004] | 0 [0 0.002] | 0.798 | 0.997 |
| Phenylalanine, median [IQR] | 0.003 [-0.013 0.014] | 0.01 [0.006 0.014] | 0.325 | 0.821 |
| Tyrosine, median [IQR] | 0.001 [-0.003 0.004] | -0.001 [-0.003 0.005] | 0.973 | 1.000 |
| Unknown signal at 7.14 ppm, median [IQR] | 0 [0 0.021] | -0.006 [-0.012 -0.003] | 0.019 | 0.238 |
| Histidine, median [IQR] | -0.003 [-0.008 0.004] | 0.001 [-0.006 0.005] | 0.812 | 0.997 |
| Glucose, median [IQR] | 0.068 [-0.113 0.665] | -0.425 [-0.48 -0.045] | 0.164 | 0.706 |
| Mannose, median [IQR] | 0.002 [-0.011 0.009] | -0.006 [-0.007 0.001] | 0.709 | 0.997 |
| Unknown signal at 5.15 ppm, median [IQR] | 0 [0 0] | 0 [0 0] | 0.057 | 0.450 |
| Unknown signal at 5.09 ppm, median [IQR] | 0 [0 0.009] | 0 [0 0] | 0.414 | 0.821 |
| Unknown signal at 5.01 ppm, median [IQR] | 0 [0 0] | 0 [0 0] | 0.904 | 0.997 |
| Ascorbate, median [IQR] | 0 [0 0.004] | -0.004 [-0.005 0] | 0.166 | 0.706 |
| Threonine, median [IQR] | 0.003 [-0.006 0.023] | 0.004 [0 0.016] | 0.919 | 0.997 |
| Lactate, median [IQR] | 0.151 [-0.193 0.375] | 0.009 [-0.719 0.046] | 0.126 | 0.644 |
| **Creatinine, median [IQR]** | **0.001 [-0.005 0.007]** | **-0.025 [-0.141 -0.004]** | **0.053** | **0.450** |
| Creatine, median [IQR] | -0.003 [-0.015 0.001] | -0.022 [-0.05 -0.003] | 0.262 | 0.785 |
| Glycine, median [IQR] | 0.022 [-0.001 0.04] | 0.026 [-0.021 0.026] | 0.292 | 0.785 |
| Methanol, median [IQR] | 0.002 [-0.001 0.018] | -0.002 [-0.002 0.001] | 0.096 | 0.544 |
| Unknown signal at 2.55 ppm, median [IQR] | 0 [0 0] | 0 [0 0] | 0.732 | 0.997 |
| Citrate, median [IQR] | -0.001 [-0.02 0.008] | -0.002 [-0.008 0.002] | 0.709 | 0.997 |
| Glutamine, median [IQR] | 0.012 [-0.019 0.04] | 0.013 [0.012 0.028] | 1.000 | 1.000 |
| Pyruvate, median [IQR] | -0.001 [-0.018 0.011] | 0.005 [-0.016 0.007] | 0.919 | 0.997 |
| Glutamate, median [IQR] | 0.034 [0.013 0.062] | 0.015 [-0.036 0.03] | 0.209 | 0.710 |
| **Acetoacetate, median [IQR]** | **0.002 [-0.032 0.018]** | **0.145 [0.074 0.259]** | **0.004** | **0.123** |
| Acetate, median [IQR] | 0.002 [-0.002 0.007] | 0.002 [-0.005 0.008] | 0.865 | 0.997 |
| Alanine, median [IQR] | 0.012 [-0.02 0.05] | 0.012 [-0.03 0.026] | 0.519 | 0.879 |
| Unknown signal at 1.45 ppm, median [IQR] | 0 [-0.004 0.009] | 0.004 [-0.008 0.012] | 0.919 | 0.997 |
| Unknown signal at 1.43 ppm, median [IQR] | 0 [0 0] | 0 [0 0] | 1.000 | 1.000 |
| **3-Hydroxybutyrate, median [IQR]** | **0.002 [-0.055 0.009]** | **0.514 [0.231 0.628]** | **0.003** | **0.123** |
| Ethanol, median [IQR] | 0 [0 0.007] | 0 [0 0] | 0.692 | 0.997 |
| Unknown signal at 1.16 ppm, median [IQR] | 0 [-0.063 0.011] | 0 [-0.895 0] | 0.534 | 0.879 |
| Unknown signal at 1.14 ppm, median [IQR] | 0 [-0.014 0.014] | 0 [-0.001 0] | 0.919 | 0.997 |
| Unknown signal at 1.11 ppm, median [IQR] | -0.004 [-0.012 0.001] | -0.003 [-0.005 -0.002] | 0.973 | 1.000 |
| Unknown signal at 1.06 ppm, median [IQR] | -0.004 [-0.007 0.003] | 0.007 [0.004 0.009] | 0.011 | 0.184 |
| Valine, median [IQR] | -0.003 [-0.015 0.028] | 0.017 [0.005 0.02] | 0.435 | 0.821 |
| Isoleucine, median [IQR] | 0 [-0.005 0.011] | 0.01 [0.003 0.012] | 0.209 | 0.710 |
| Leucine, median [IQR] | 0 [-0.007 0.012] | 0.008 [0.003 0.01] | 0.359 | 0.821 |
| 2-Hydroxybutyrate, median [IQR] | -0.006 [-0.02 0] | -0.003 [-0.007 0.004] | 0.395 | 0.821 |
| Protein NH, median [IQR] | 2.231 [-6.081 10.547] | -3.854 [-6.531 -2.663] | 0.209 | 0.710 |
| Unsaturated lipid -CH=CH-, median [IQR] | -0.154 [-0.404 1.137] | 0.667 [0.489 0.905] | 0.519 | 0.879 |
| Lipid alpha-CH2, median [IQR] | -0.03 [-0.286 0.1] | 0.344 [-0.166 0.492] | 0.476 | 0.866 |
| Cholesterol, median [IQR] | 0.005 [-0.052 0.066] | 0.049 [0.038 0.148] | 0.083 | 0.530 |
| Lipid =CH-CH2-CH=, median [IQR] | -0.01 [-0.139 0.513] | 0.232 [0.177 0.469] | 0.292 | 0.785 |
| Glycorol phospholipid, median [IQR] | -0.028 [-0.139 0.02] | 0 [-0.332 0.051] | 0.865 | 0.997 |
| Phospholipid, median [IQR] | 0.031 [-0.164 0.214] | 0.06 [-0.077 0.061] | 0.760 | 0.997 |
| Lipid beta-CH2, median [IQR] | -0.111 [-0.267 0.472] | 0.386 [0.025 0.453] | 0.396 | 0.821 |
| Lipid CH2, median [IQR] | -0.403 [-3.014 3.979] | 2.495 [0.349 4.314] | 0.564 | 0.898 |
| Lipid CH3, median [IQR] | 0.366 [-0.619 1.49] | 0.679 [0.518 1.528] | 0.435 | 0.821 |
| GlycB, median [IQR] | 0.035 [0.002 0.049] | 0.014 [-0.015 0.026] | 0.262 | 0.785 |
| GlycA, median [IQR] | 0.126 [0.009 0.267] | -0.045 [-0.068 0.01] | 0.062 | 0.450 |

FDR – False discovery rate, IQR- Interquartile range

**Table S12.** Comparison of metabolites with organ dysfunction in acute pancreatitis. Significantly dysregulated lipids and other significantly altered metabolites are represented in bold.

| **Feature** | **FALSE** | **TRUE** | **p-value** | **FDR** |
| --- | --- | --- | --- | --- |
| Formate, median [IQR] | 0.002 [0 0.004] | 0.003 [-0.002 0.004] | 0.978 | 0.997 |
| Unknown signal at 8.12 ppm, median [IQR] | 0.002 [-0.001 0.005] | 0.001 [-0.001 0.006] | 0.637 | 0.792 |
| Unknown signal at 8.07 ppm, median [IQR] | 0 [0 0] | 0.001 [0 0.003] | 0.113 | 0.339 |
| Phenylalanine, median [IQR] | -0.005 [-0.014 0.013] | 0.008 [0.003 0.014] | 0.212 | 0.491 |
| Tyrosine, median [IQR] | 0.001 [-0.003 0.005] | 0 [-0.003 0.004] | 0.718 | 0.852 |
| Unknown signal at 7.14 ppm, median [IQR] | 0 [-0.001 0.021] | 0 [-0.005 0.013] | 0.597 | 0.780 |
| Histidine, median [IQR] | -0.005 [-0.008 0.003] | 0.004 [-0.003 0.006] | 0.063 | 0.248 |
| Glucose, median [IQR] | -0.053 [-0.291 0.266] | 0.068 [-0.33 0.502] | 0.524 | 0.722 |
| Mannose, median [IQR] | -0.001 [-0.014 0.005] | 0.002 [-0.007 0.012] | 0.332 | 0.604 |
| Unknown signal at 5.15 ppm, median [IQR] | 0 [0 0] | 0 [0 0] | 0.462 | 0.722 |
| Unknown signal at 5.09 ppm, median [IQR] | 0 [-0.001 0.001] | 0.003 [0 0.008] | 0.283 | 0.535 |
| Unknown signal at 5.01 ppm, median [IQR] | 0 [0 0] | 0 [0 0] | 0.492 | 0.722 |
| Ascorbate, median [IQR] | 0 [0 0.004] | 0 [-0.005 0] | 0.192 | 0.466 |
| Threonine, median [IQR] | 0.004 [-0.006 0.029] | 0.003 [-0.003 0.014] | 0.978 | 0.997 |
| Lactate, median [IQR] | 0.019 [-0.197 0.365] | 0.068 [-0.15 0.158] | 1.000 | 1.000 |
| Creatinine, median [IQR] | -0.003 [-0.011 0.005] | 0 [-0.007 0.007] | 0.978 | 0.997 |
| Creatine, median [IQR] | -0.004 [-0.017 0] | -0.003 [-0.021 0] | 0.934 | 0.997 |
| Glycine, median [IQR] | 0.013 [-0.003 0.028] | 0.035 [0.026 0.045] | 0.081 | 0.257 |
| Methanol, median [IQR] | 0.002 [-0.005 0.029] | 0.001 [-0.001 0.002] | 0.524 | 0.722 |
| Unknown signal at 2.55 ppm, median [IQR] | 0 [0 0] | 0.004 [0 0.058] | 0.027 | 0.140 |
| Citrate, median [IQR] | -0.007 [-0.016 0.002] | 0.001 [-0.015 0.008] | 0.421 | 0.693 |
| Glutamine, median [IQR] | -0.003 [-0.043 0.043] | 0.019 [0.012 0.028] | 0.390 | 0.663 |
| Pyruvate, median [IQR] | -0.01 [-0.022 0.002] | 0.006 [0 0.022] | 0.071 | 0.257 |
| Glutamate, median [IQR] | 0.025 [0 0.05] | 0.029 [0.018 0.063] | 0.524 | 0.722 |
| **Acetoacetate, median [IQR]** | **0.001 [-0.022 0.008]** | **0.094 [0.043 0.231]** | **0.007** | **0.121** |
| Acetate, median [IQR] | 0 [-0.003 0.007] | 0.003 [0 0.005] | 0.978 | 0.997 |
| Alanine, median [IQR] | 0.015 [-0.032 0.05] | 0.002 [-0.014 0.027] | 0.637 | 0.792 |
| Unknown signal at 1.45 ppm, median [IQR] | 0 [-0.005 0.006] | 0.002 [-0.005 0.014] | 0.524 | 0.722 |
| Unknown signal at 1.43 ppm, median [IQR] | 0 [0 0] | 0 [0 0] | 0.175 | 0.447 |
| **3-Hydroxybutyrate, median [IQR]** | **0.001 [-0.046 0.005]** | **0.275 [0.033 0.599]** | **0.011** | **0.134** |
| Ethanol, median [IQR] | 0 [0 0.009] | 0 [0 0] | 0.264 | 0.518 |
| Unknown signal at 1.16 ppm, median [IQR] | 0 [-0.082 0.023] | 0 [-0.055 0] | 0.652 | 0.792 |
| Unknown signal at 1.14 ppm, median [IQR] | 0 [-0.011 0.017] | 0 [-0.003 0.004] | 0.889 | 0.997 |
| Unknown signal at 1.11 ppm, median [IQR] | -0.01 [-0.014 -0.002] | -0.002 [-0.004 0.003] | 0.142 | 0.380 |
| Unknown signal at 1.06 ppm, median [IQR] | -0.003 [-0.006 0.003] | 0.003 [-0.006 0.008] | 0.255 | 0.518 |
| Valine, median [IQR] | -0.003 [-0.02 0.014] | 0.019 [-0.003 0.045] | 0.127 | 0.360 |
| Isoleucine, median [IQR] | 0 [-0.007 0.01] | 0.011 [0.002 0.014] | 0.081 | 0.257 |
| **Leucine, median [IQR]** | **-0.002 [-0.008 0.007]** | **0.009 [0.002 0.02]** | **0.038** | **0.159** |
| 2-Hydroxybutyrate, median [IQR] | -0.007 [-0.015 0] | -0.003 [-0.016 0] | 0.560 | 0.751 |
| Protein NH, median [IQR] | 1.147 [-6.205 6.871] | -1.031 [-5.977 7.473] | 0.890 | 0.997 |
| **Unsaturated lipid -CH=CH-, median [IQR]** | **-0.247 [-0.625 0.088]** | **0.786 [0.505 1.611]** | **0.018** | **0.134** |
| **Lipid alpha-CH2, median [IQR]** | **-0.271 [-0.361 0.03]** | **0.418 [0.045 0.603]** | **0.002** | **0.106** |
| **Cholesterol, median [IQR]** | **-0.004 [-0.061 0.039]** | **0.052 [0.03 0.156]** | **0.021** | **0.136** |
| **Lipid =CH-CH2-CH=, median [IQR]** | **-0.113 [-0.215 0.051]** | **0.351 [0.151 0.636]** | **0.018** | **0.134** |
| **Glycorol phospholipid, median [IQR]** | **-0.11 [-0.279 0]** | **0.033 [-0.002 0.138]** | **0.025** | **0.140** |
| Phospholipid, median [IQR] | -0.003 [-0.313 0.135] | 0.063 [-0.008 0.206] | 0.255 | 0.518 |
| **Lipid beta-CH2, median [IQR]** | **-0.149 [-0.514 0.041]** | **0.42 [0.112 1.023]** | **0.014** | **0.134** |
| **Lipid CH2, median [IQR]** | **-2.623 [-5.925 0.397]** | **5.321 [0.886 9.455]** | **0.007** | **0.121** |
| **Lipid CH3, median [IQR]** | **0.117 [-1.375 0.627]** | **1.104 [0.486 2.753]** | **0.038** | **0.159** |
| GlycB, median [IQR] | 0.012 [0 0.037] | 0.042 [0.028 0.056] | 0.233 | 0.517 |
| GlycA, median [IQR] | 0.019 [-0.034 0.18] | 0.132 [0.037 0.279] | 0.360 | 0.633 |

FDR – False discovery rate, IQR- Interquartile range
